# Stacking multiple prediction models to optimise performance in local settings: exemplars in cardiometabolic disease

**DOI:** 10.1101/2023.06.16.23291489

**Authors:** Sreejita Ghosh, Jasmine Gratton, Roel Vermeulen, Folkert Asselbergs, Jelle J. Vlaanderen, A. Floriaan Schmidt

## Abstract

**Background:** Risk prediction models are used in healthcare settings to tailor therapies to individuals most likely to benefit. Despite appropriate external validation, difference in local characteristics (e.g. patient mix) may attenuate model performance. Prior to any implementation it is therefore advisable to explore local performance, typically requiring a modest amount of historic data. Depending on model performance, model adjustments might be necessary which often require large amounts of data. Here we explore a small sample size approach approximating de novo derivation, by combining model stacking and transfer learning, referred to as *stacked transfer learning*. As an example we focus on stacking previously trained risk prediction models for cardiovascular disease (CVD), stroke, (chronic) kidney disease, and diabetes.

**Methods:** We leverage data from the UK biobank to illustrate the benefits of stacking previously trained risk prediction models, predicting the risk of incident CVD, chronic kidney disease (CKD) or diabetes. To mimic sample sizes available in local settings, such as a small to large healthcare trust, we iterated the number of training cases between 10 and 1000. Model stacking was performed using a LASSO penalized logistic regression model, and compared performance of a *de novo* model estimating the local association of 33 variables used in the aforementioned risk prediction models.

**Results:** We found that stacked models require roughly one-tenths of the training sample size compared to de novo derivation of a prediction model. For example, predicting CVD the stacked model required 30 cases to reach a area under the curve (AUC) value (with 95% CI) of 0.732 (0.728, 0.735), while the *de novo* model required 300 cases to reach approximately the same performance. As expected, the absolute performance depended on the predicted outcome, where for example the difference between *de novo* and stacked modelling was smaller for CKD prediction.

**Conclusion:** We show that our proposed ”stacked transfer learning” approach closely approximated the predictive performance of a *de novo* model, often requiring only a fraction of the data. As such, this approach should be considered when tailoring a model to a local setting.

## INTRODUCTION

In clinical settings, risk prediction models are often employed to tailor treatment to people at risk of future disease or disease progression. For cardiovascular disease (CVD) (a composite of coronary heart disease (CHD) and stroke), risk-stratified initiation of treatments is recommended by professional bodies such as the American College of Cardiology/American Heart Association (ACC/AHA) [1] and the European Society of Cardiology (ESC)[2, 3]. The UK National Institute for Health and Care Excellence (NICE) suggest using the QRISK2 or QRISK3 algorithms[4, 5, 6, 7] and recommend pharmaceutical interventions for people with a 10-year predicted risk of 10% or higher [8].

Because of the importance of these tools for clinical disease management, *local* (e.g., such as in a local healthcare trust) model under-performance may have detrimental effects, where low-risk people may be over-treated, and high-risk people may not receive timely treatment. While some of these risk prediction models, including QRISK3, have been externally validated, local performance may differ substantially. For example, the SCORE2 model [9] for subsequent CVD events has been extensively validated across Europe, showing variable discriminative ability, where the area under Receiver Operating Characteristics (ROC) curve (AUC) ranged between 0.67 (in Russia) and 0.83 (in Italy).

As such, and notwithstanding promising performance in external setting, risk prediction models may under-perform in a new settings. Before considering model implementation, it is therefore advisable to first explore local performance. Depending on this local performance, a model might need to be tailored to improve its performance, which could be achieved through the principles of transfer learning[10]. For example, calibration (the agreement between predicted and observed risk) can often be improved through a simple re-calibration step, estimating a new intercept (reflecting the average disease risk) and calibration slope; which is equivalent to vertically translating and scaling the predicted risk.

More data-intensive calibration steps include adding additional predictor variables or candidate features, or even de novo derivation of an entire prediction model [11]. Typically these more elaborate re-calibration steps require a sample size unavailable in even large healthcare trusts. Instead, we propose to leverage the large body of already available pre-trained prediction models[12], and use the principles of model *stacking*[13] and *transfer learning*[14, 15], to combine a number of already derived prediction models to jointly predict disease risk. We will show that such *stacked transfer learning* will require a fraction of the sample size necessary to train a *de novo* model using all available predictors, and as such might be realistically applied to optimize risk prediction models in local settings. Furthermore, because such a stacked transfer learning approach often approximates performance of a de novo modelling strategy (at smaller sample sizes), this provides a bench-mark to compare alternative modelling steps (e.g., no-updating or updating a small number of predictors).

As an exemplar, we attempted to predict 10-year CVD risk in the UK biobank (UKB) by stacking four prediction models derived by QResearch: QRISK3 for CVD [7], Q-Stroke for stroke [16], Q-Diabetes for type 2 diabetes (T2D) [17], or Q-Kidney for chronic kidney disease (Q-CKD). Performance of these stacked models was compared to the original QRISK3 (as well as the four other Q-scores), and to a (*de novo*) model (referred to as *de novo* hereafter) trained to predict CVD using all of the individual predictors used in at least one of the Q-scores. To mimic the amount of samples likely available in real-world settings, we performed an empirical simulation study and took random samples from a training split of the UKB to create a range of cohorts with different sample sizes (including 10 to 1000 CVD cases). This empirical simulation study was repeated for the stacked and the *de novo* model attempting to predict the 10-year risk of T2D and the 5-year risk of CKD.

## METHODS

### Data source

Data were available from UKB participants including information from hospital episode statistics (HES) and general practitioners (GP). Participants were enrolled between 2006 and 2010 from across the UK.

To empirically showcase the concept of transfer learning using stacked models we set out to predict the 10-year risk of incident CVD in people without a history of CHD, atrial fibrillation, or heart failure. Incident CVD was defined based on the ICD-10 codes listed in Appendix Table A1. Similarly we explored the predictive performance of the 10-year risk of T2DM, and 5-year risk of CKD, in people without a history of these respective diseases at enrolment. Supplementary Table A2 details the candidate features used by the four Q-scores. Briefly the following data were extracted for each UKB participants at the time of enrolment: Female sex, age (years), body mass index (BMI), high-density lipoprotein cholesterol (HDL-C, mmol/L), low-density lipoprotein cholesterol (LDL-C, mmol/L), triglycerides (LDLC, mmol/L), total cholesterol (mmol/L), systolic blood pressure (SBP, mm Hg), smoking status (never, former, light, moderate, and heavy), and disease histories. Prescription data were extracted for the following drugs: statins, corticosteroids, non-steroidal anti-inflammatory drugs (NSAIDs), and atypical antipsychotics.

The limited number of missing observations (Table 1, A6 and A5) were imputed using multivariate imputation by chained equations (MICE) [18].

**Table 1.** Baseline characteristics of UK biobank participants without CVD at the time of enrolment.

| Participant characteristics | Cases | Controls | Missing-ness % |
| --- | --- | --- | --- |
| Total sample size (n) | 23389 | 302265 |  |
| Age (median [IQR]) | 63.0 (58.0 , 66.0) | 57.0 (50.0 , 63.0) | 0.0 |
| Sex (male) (%) | 14763.0 (63.12%) | 131364.0 (43.46%) | 0.0 |
| Systolic blood pressure in mmHg (median [IQR]) | 145.0 (133.0 , 159.0) | 138.0 (126.0 , 152.0) | 0.09 |
| HDL cholesterol in mmol/L (median [IQR]) | 1.3 (1.1 , 1.55) | 1.42 (1.19 , 1.7) | 12.74 |
| LDL cholesterol in mmol/L (median [IQR]) | 3.48 (2.84 , 4.13) | 3.57 (3.02 , 4.16) | 4.83 |
| Triglycerides in mmol/L (median [IQR]) | 1.68 (1.19 , 2.41) | 1.47 (1.04 , 2.13) | 4.73 |
| BMI in kg/m2 (median [IQR]) | 27.95 (25.22 , 31.3) | 26.53 (23.99 , 29.6) | 0.30 |
| Smoking status (%) |  |  | 0.04 |
| Non-smoker | 10375 ( 44.0 %) | 169905 (56.0%) |  |
| Former smoker | 9711 ( 42.0 %) | 102393 ( 34.0 %) |  |
| Light smoker (< 10 cigarettes/day) | 1012 ( 4.0 %) | 8359 ( 3.0 %) |  |
| Moderate smoker (10-19 cigarettes/day) | 925 ( 4.0 %) | 7171 ( 2.0 %) |  |
| Heavy smoker (> 20 cigarettes/day) | 300 ( 1.0 %) | 4011 ( 1.0 %) |  |
| Diagnosed/ treated hypertension | 2505.0 (10.71%) | 11579.0 (3.83%) | 0.0 |
| History of Type 1 Diabetes | 39.0 (0.17%) | 401.0 (0.13%) | 0.0 |
| History of Type 2 Diabetes | 1178.0 (5.04%) | 5764.0 (1.91%) | 0.0 |
| History of atrial fibrillation | 0.0 (0.0%) | 0.0 (0.0%) | 0.0 |
| History of peripheral vascular disease | 169.0 (0.72%) | 496.0 (0.16%) | 0.0 |
| History of valvular heart disease | 194.0 (0.83%) | 403.0 (0.13%) | 0.0 |
| History of chronic heart disease | 0.0 (0.0%) | 0.0 (0.0%) | 0.0 |
| History of congestive cardiac failure | 0.0 (0.0%) | 0.0 (0.0%) | 0.0 |
| History of rheumatic arthritis | 197.0 (0.84%) | 1086.0 (0.36%) | 0.0 |
| History of Systemic lupus erythematosus | 25.0 (0.11%) | 146.0 (0.05%) | 0.0 |
| History of chronic kidney disease | 84.0 (0.36%) | 232.0 (0.08%) | 0.0 |
| History of end stage kidney disease | 84.0 (0.36%) | 232.0 (0.08%) | 0.0 |
| History of kidney stones | 172.0 (0.74%) | 1187.0 (0.39%) | 0.0 |
| History of migraine | 184.0 (0.79%) | 2472.0 (0.82%) | 0.0 |
| History of severe mental illness | 114.0 (0.49%) | 1054.0 (0.35%) | 0.0 |
| Prescription history of Statins | 5465.0 (23.37%) | 31850.0 (10.54%) | 0.0 |
| Prescription history of atypical antipsychotics | 76.0 (0.32%) | 800.0 (0.26%) | 0.0 |
| Prescription history of corticosteroids | 668.0 (2.86%) | 4355.0 (1.44%) | 0.0 |
| Prescription history of NSAID | 8378.0 (35.82%) | 73234.0 (24.23%) | 0.0 |

### Implementation of the Q-scores

The Q-scores were implemented using the publicly available code shared by the QResearch group. Based on known shared etiology we focused on implementing the aforementioned scores for CVD (QRISK3), stroke (Q-Stroke), T2D (Q-Diabetes), and for CKD (Q-CKD). Please see the appendix methods for a full description of the implementation; also noting some necessary simplification, for example, due to the lack of repeat SBP measurements, we implemented a version of the QRISK3 which omitted the standard deviation (SD) of SBP measurements.

### Stacked transfer learning

In machine learning (ML), transfer learning refers to adapting a pre-trained model to perform a new but related task[19, 14]. For example, an algorithm that is trained to identify and extract left ventricle ejection fraction from medical notes can be repurposed to extract information on additional measurements sourced for example from a different healthcare trust. By sourcing previously obtained knowledge and updating it to local settings, transfer learning minimizes the amount of data needed to obtain a model that is sufficiently accurate for the new task [15]. Here we propose a specific type of transfer learning, leveraging and combining information from previously trained risk prediction models to obtain a locally optimized algorithm. In the context of transfer learning, combining (or stacking) multiple previously derived prediction models can be viewed as a weighted sum of the predicted risk of each individual model that might explain the outcome (Figure 1), where the weights need to be estimated from a set of historical data. For example, stacked transfer learning can be implemented as a relatively straightforward logistic regression model only considering linear terms:

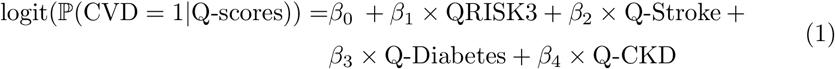

While the individual Q-scores include 16 to 22 candidate features, the linear stacked model presented in equation 1 instead contains only four features which typically can be estimated with data from fewer participants than what is needed for fitting the *de novo* model with 33 candidate features.

**Figure 1.**
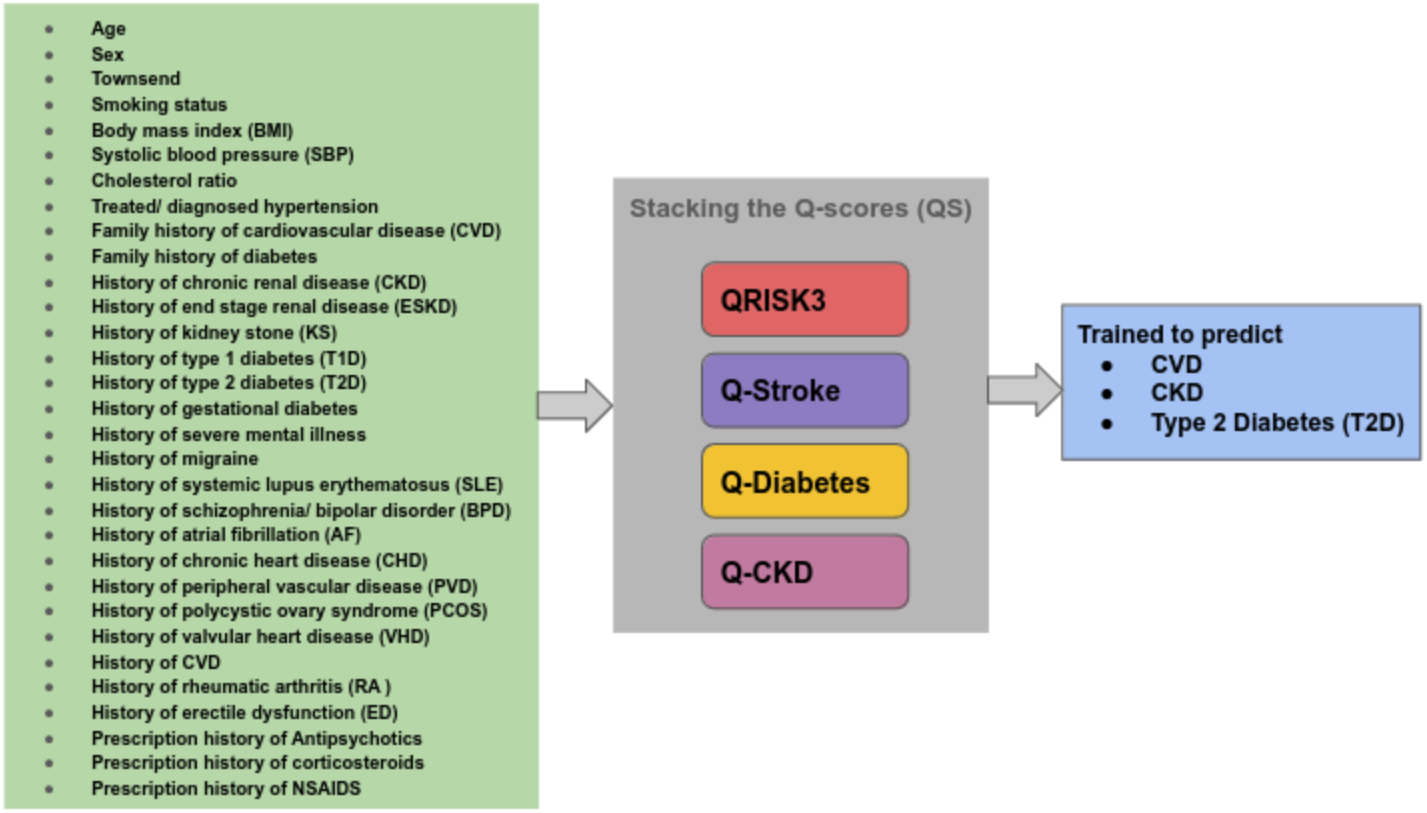
Schematic of our proposed stacked transfer learning approach applied to the Q-research risk equations for the onset of 10-years CVD, 10-years T2D and 5-years CKD.

Model performance may be further improved by considering more complex non-linear terms (e.g. squared terms or product terms). For example a three-way polynomial of the four Q-scores would result in 35 candidate predictors (including the terms with higher powers), which can be contrasted by a three-way polynomial of the 33 feature *de novo* model resulting in 5456 candidate predictors. Given the large number of candidate predictors, model selection strategies such as LASSO penalization can be employed to identify the subset of predictors which may optimally predict the outcome.

Depending on the desired model flexibility, the LASSO model can be substituted by more flexible algorithms, for example a random forest [20, 21] or support vector machines [22, 23]. In the current manuscript we consider the two types of stacked models (1) a linear model equal to Equation 1 (*stacked: linear*) and (2) a 3-way polynomial model (*stacked: 3-way poly*). Here both models were implemented using a binomial generalized linear model, with LASSO penalization optimized through 5-fold cross-validation. As a comparison, we used the same generalized linear model with LASSO regularization to train a *de novo* model using 33 candidate features; see Table A2.

### Performance across different sample sizes

Given that in most local settings a sample size of 300K+ participants is unfeasible, and to ensure unbiased performance evaluation, we split the UKB into a 85/15% testing and training set. To empirically simulate model performance across varying sample size and case number, we iterated from case numbers as low as 10 to 100 in steps of 10, and further until 1000 in steps of 100. Random draws were taken exclusively from the training data, stratified by case-control status, ensuring a constant event rate.

Additionally, for the models predicting CVD incidence within a period of 10 years, we compared the performance of the stacked and *de novo* models to each individual Q-score using a representative sample size of 100 cases (resulting in a total sample size of 1384).

### Discrimination measures and calibration estimates

In this study, external performance (using the test data) was evaluated in terms of discrimination ability (AUC) and calibration (calibration slope and calibration-in-the-large), with precision indicated using 95% confidence intervals (95%CI). Additionally, using the test data, feature importance was estimated by sequential permutation (25 times) of each individual predictor variable and recording the change in AUC.

Perfect discrimination is indicated by an AUC of 1.00, where 0.50 reflects random (or no) discrimination. Calibration indicates how accurate the absolute predicted risk is, where calibration-in-the-large reflects the average difference between observed and predicted risk (on the logit scale) and the calibration slope reflects potential deviations in the tails of the distribution [11, 24, 25]. Hence perfect calibration is obtained when calibration-in-the-large is 0 and the calibration slope is 1 [26, 27].

## RESULTS

### Baseline characteristics

During a 10-year follow-up, 23,389 CVD events were accrued out of 325,654 UKB participants without a history of CVD at enrolment (Table 1). At the time of enrolment, participants (both cases and control)) had a median age of 56.6 years (SD: 8.00), 179,527 (55.12%) were female, and the average BMI was 27.303 (SD: 4.72). Mean LDL-C concentration was 3.606 mmol/L (SD: 0.856), mean HDL-C concentration was 1.463 mmol/L (SD: 0.381), 37,315 (11.5 %) participants had a statin prescription, and the average SBP was 140.2 mmHg (SD: 19.643). Please see Table A6 and Table A5 for the baseline characteristics of the CKD and T2D cohorts.

Given that the Q-scores partially included the same information (e.g., they all used age and sex as predictors), we assessed the pairwise correlation of variables. Aside from a meaningful correlation between Q-CVD and Q-Stroke (Spearman’s correlation of 0.799), the Q-research scores were only moderately correlated.

### Evaluating the influence of case numbers for CVD incidence prediction

We first compared performance of the two transfer learning models to the *de novo* model across different amounts of training data that one might expect in a reasonable clinical (non-research) setting (by iterating the number of cases while keeping the incidence proportion constant) (Figure 2). The stacked transfer learning models consistently showed a larger AUC than the *de novo* model for 30 to 400 cases. Figure 2 also shows that the AUC values of the stacked Q-score models almost overlap across the range of case numbers investigated, indicating a lack of non-linear associations; Table A3. The calibration (i.e., the agreement between predicted risk and observed risk) was near acceptable for both stacked model and the *de novo* model from a case number of 90 onward; see Figure 3 .

**Figure 2.**
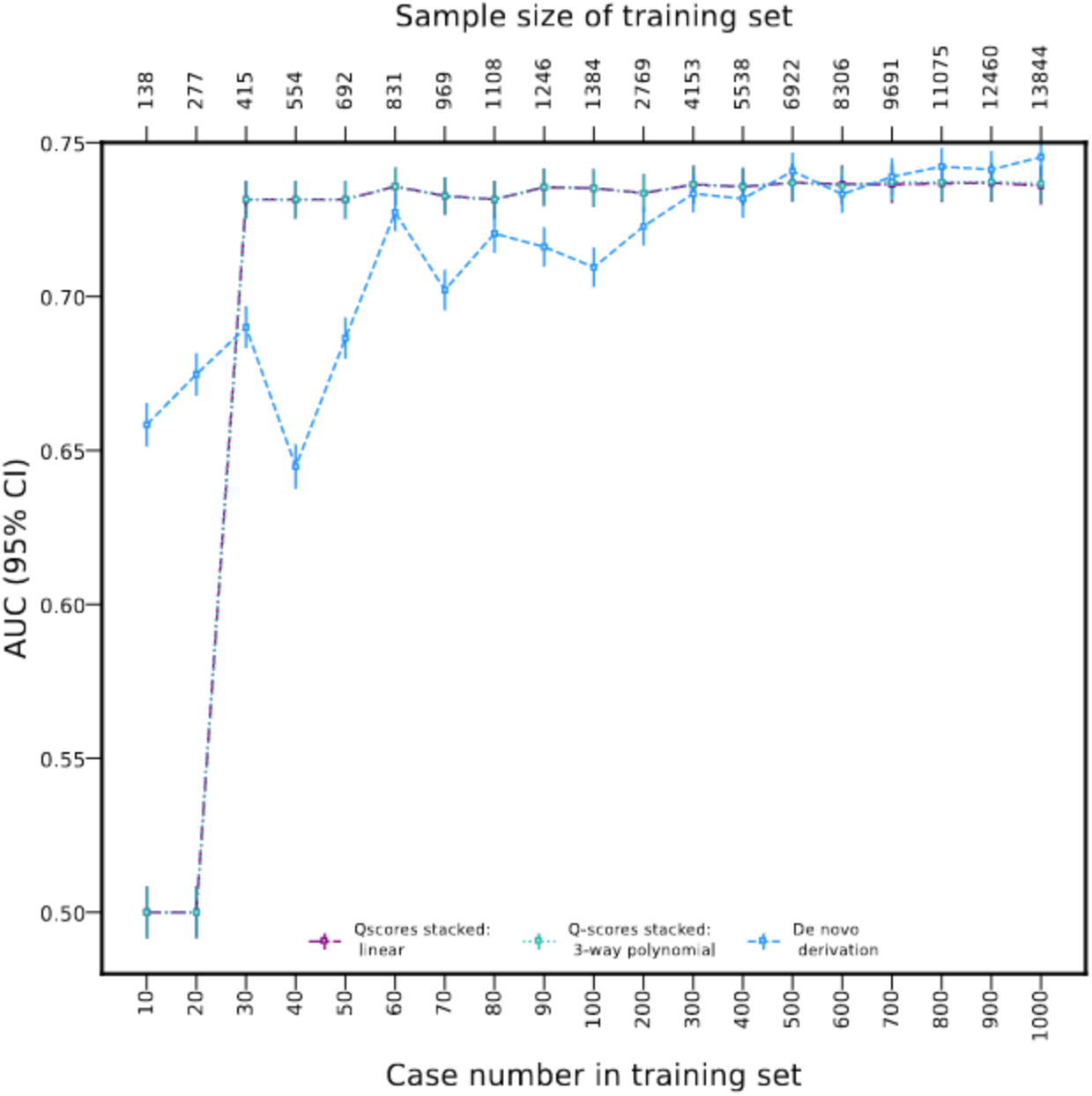
Model performance across a range of training sample sizes possibly encountered in non-research healthcare settings. Performance of the two stacked models overlap across this given range. Estimates were derived from an independent test set of 18,849 CVD cases and a total of 276,646 participants. The numerical data underpinning this figure are provided in Table A3.

**Figure 3.**
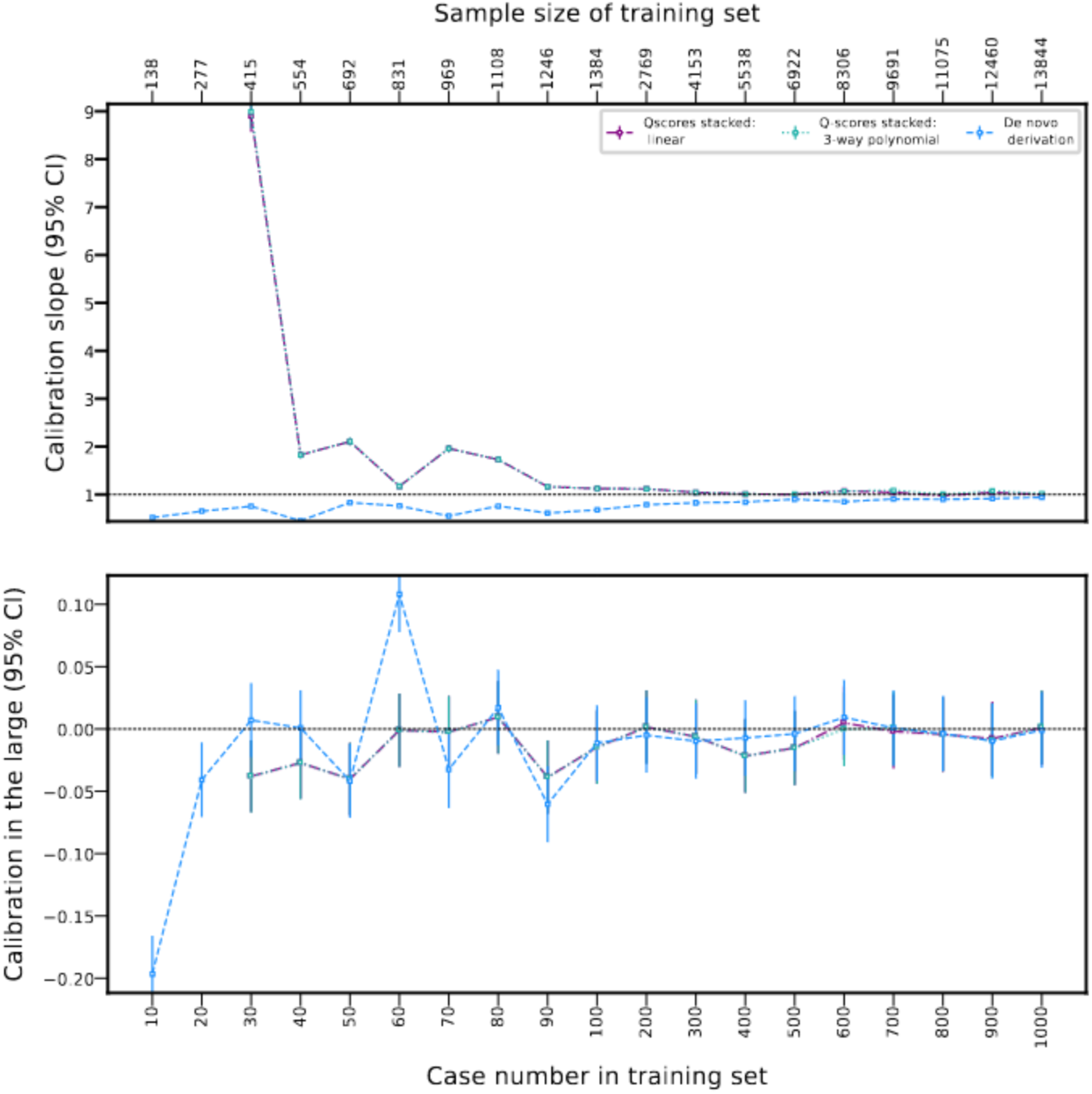
Plots of the calibration estimates, illustrating the agreement between predicted CVD risk and observed CVD risks for the stacked Q-risk score models and the *de novo* model, across a range of training sample sizes and corresponding case numbers. Estimates were derived from an independent test set of 18,849 CVD cases and a total of 276,646 participants. The numerical data underpinning this figure are provided in Table A3.

### Comparing the stacked and the *de novo* models, against individual Q-scores

Next we studied the performance of the stacked and *de novo* models against individual Q-scores for a representative example of 100 CVD events (total sample size = 1384). Figure 4 and Table A4 showed that among the Q-scores, QRISK3 reached the highest AUC 0.732 (95%CI 0.729; 0.735), followed by Q-CKD 0.692 (95%CI 0.688; 0.695), Q-Stroke 0.689 (95%CI 0.686; 0.693), and Q-Diabetes 0.674 (95%CI 0.671; 0.678). The stacked models showed a slightly improved discriminative ability: AUC 0.735 (95%CI 0.732; 0.738) for both the linear and 3-way polynomial stacked models. For this same sample size, the discriminative ability (AUC) of the *de novo* model was 0.710 (95% 0.706; 0.713). At this sample size, the stacked model were better calibrated (calibration-in-the-large of -0.015 (95 % CI -0.029, 0.000)) than the *de novo* model(calibration-in-the-large of -0.011 (95% CI -0.026, 0.004)), also outperforming the QRISK3 (calibration-in-the-large of -0.308 (95% CI -0.323, -0.294)).

**Figure 4.**
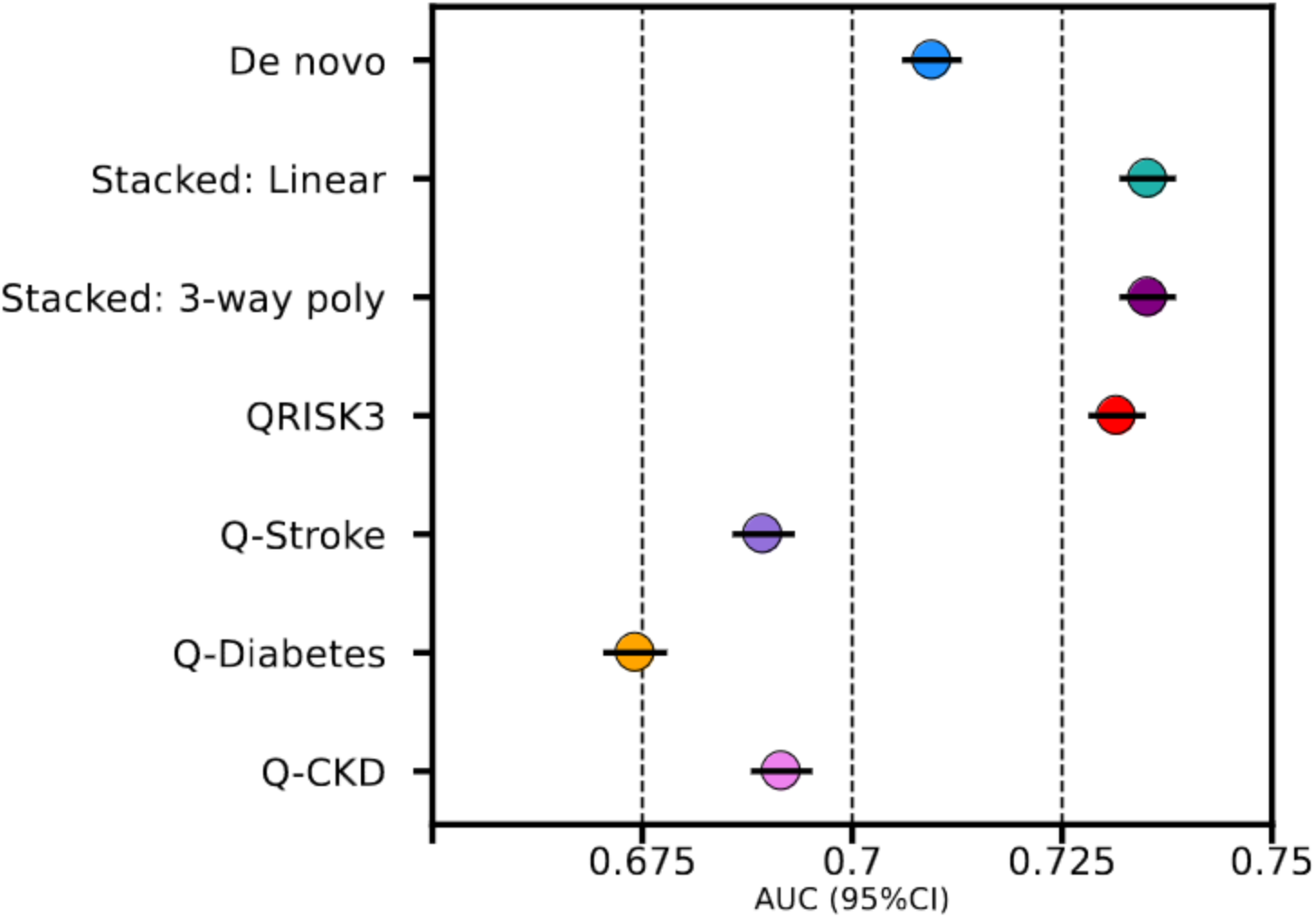
The discriminative ability of various prediction models to identify people who experienced a CVD event during a 10-year follow-up period. N.B. Discrimination was estimated as the AUC with 95% confidence intervals (CI). The stacked and *de novo* models were trained on an illustrative example dataset of 100 CVD cases (1384 samples in total). Performance was evaluated in a hold-out test set of 19,849 cases and a total sample size of 276,646. The numerical data underpinning this figure are provided in Table A4. Please see Figure 2 and 3, and Table A3 for performance of the entire range of sample sizes.

### Predicting 10-year and 5-year risk of type 2 diabetes and chronic kidney disease respectively

To further explore the utility of our strategy of stacked transfer learning, we used these same four Q-scores to predict the incidence of T2D or CKD over a similar 10-year period (for T2D) or 5-year period (for CKD).

In both examples (Figure 5), the stacked models showed comparable performance with respect to that of the *de novo* model, even at relatively low case numbers. In fact at low case numbers (therefore low sample sizes) the stacked models were generally better calibrated compared to the *de novo* model, Figure A5 and A6, Table A7 and A8. Figure A4 and Figure A3 rank the predictors in the *denovo* model, and the four Q-scores in the stacked models, in order of their contributions to these models for the prediction of incident CKD and T2D respectively.

**Figure 5.**
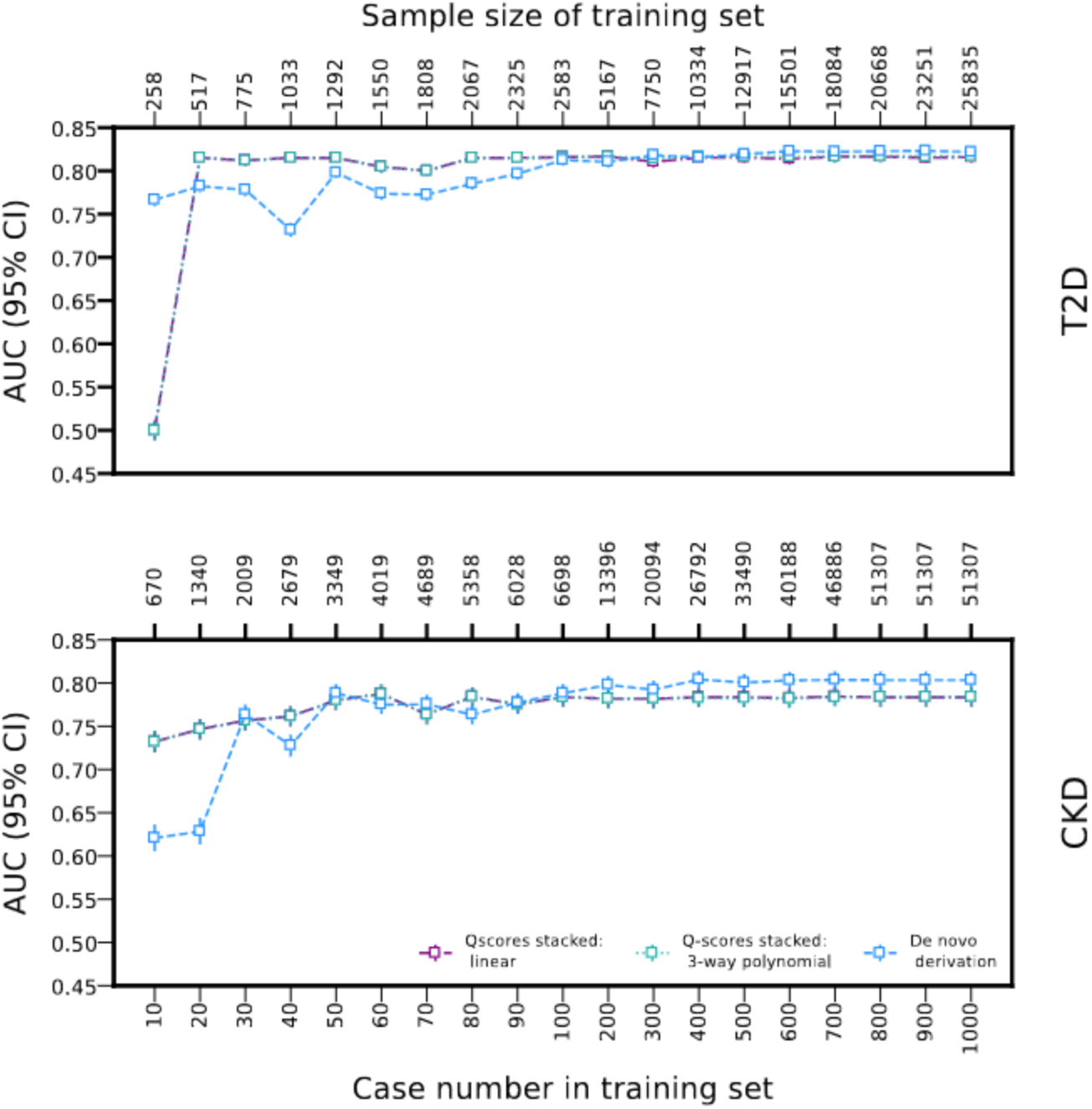
Model performance across a range of training sample sizes that could be encountered in non-research healthcare settings, for predicting the 10-year risk of T2D and the 5-year risk of CKD. AUC estimates were derived from an independent test set of (a) 10,683 T2D cases from a total of 279,290 participants, and (b) 4,390 CKD cases from a total of 289,594 participants.The numerical data underpinning this figure are provided in Table A7 and A8.

The general agreement between the stacked and *de novo* models in terms of discrimination was explored through permuted feature importance, which highlighted that while for predicting outcome of incident T2D, Q-Diabetes was predominant with minor contribution from QRISK3, for predicting the outcome of incident CKD, it was the combined contribution of QRISK3, Q-Diabetes along with Q-CKD. This suggests benefits of stacking multiple phenotypically distinct models.

## DISCUSSION

In this paper we demonstrated how the wealth of pre-trained prediction models can be combined to derive a locally optimized model in small sample size settings (or equivalently in settings a small number of cases). We refer to this approach as ”stacked transfer learning”. First we used CVD incidence prediction as an empirical example to show that this approach can approximate the performance of a *de novo* model at considerably lower sample size settings (e.g, using only 10% of the number of cases necessary for the de novo derivation model). Using additional examples in T2D and CKD prediction, we con firmed similar performance against deriving a *de novo* model.

As an illustrative example we considered the 10-year prediction of incident CVD. Due to the availability of guideline-recommended risk-stratified treatment initiation and escalation[8], a large number of CVD prediction rules have been derived, and while some of these models have been externally validated, it is widely appreciated that performance will vary between local settings[12]. Conditional on acceptable performance in independent test data (providing an unbiased estimate of model performance in the same type of data), difference in model performance between distinct local settings may reflect changes in ”case-mix”[28]. For example, over time, one would expect medical care to improve, impacting the patient characteristics (i.e., presenting with different co-morbidities and/or co-prescriptions). Difference in country or more general the type of care (i.e., primary or secondary care) will additionally impact performance, and may require calibrating a model to local settings. Therefore, as an alternative to creating setting-specific algorithm, we propose that tailoring a ”base-line” model (which has been extensively validated already) to accommodate setting-specific particularities might provide a more uniform approach. We show that our approach of stacked transfer learning might be a relatively straightforward tool for this.

For illustrative purposes we implemented our stacked transfer learning approach using fairly standard binomial generalized linear models with a LASSO penalization. Depending on the desired model complexity (i.e., the need to accommodate non-linear or interaction effect), alternative supervised models may be used as ”drop-in-replacements”, for example (regularized) support vector machine [22, 23] might provide additional modelling flexibility without requiring vasts amounts of participants. It is worth nothing that in our empirical example, including non-linear and interaction terms did not meaningfully impact performance.

While a great number of CVD risk prediction models have been derived, as we showed here, our stacked learning approach does not presuppose to only combine models which were originally trained to predict CVD. Given that the various models are stacked (i.e., combined) as a weighted average, where the weights are empirically derived using a supervised method, models which do not meaningfully explain the outcome risk will simply get a smaller weight (or depending on the type of penalization, a zero weight). For example, in our analysis, the Q-CKD of the stacked models had a feature importance close to zero indicating that it contributed little to the 10-year CVD prediction risk (relative to the other 3 scores). An understandable objection to combining predicted risk from multiple pre-trained models is a subset of the input data (e.g., age, sex, BMI) is shared between the models. However, unless the predicted risk is highly dependent on this subset of variables, it is unlikely that this would result in meaningful correlation which might lead to model instability (as shown in our correlation analysis in Figure A1), and such cases can be readily identified and addressed analytically (e.g., leave a model out or increase penalization). This is demonstrated in Figure 4 and A2, which illustrate that even though among the individual Q-scores, Q-Diabetes had the lowest AUC for predicting the CVD outcome, the second highest contribution (feature importance) in both the stacked models comes from Q-Diabetes itself. Conversely, while contribution of Q-CKD to the stacked models is close to zero, the AUC of Q-CKD for predicting the CVD outcome is second highest after QRISK3. Similarly, while we focused on stacking predicted risk from pre-trained models, this approach can readily include additional candidate predictor variables relevant for a local setting (e.g., the presence or absence of familial hypercholesterolemia carriership).

The following limitations deserve consideration. While the principles of stacked transfer learning follow directly from first principle, we have only provided limited empirical simulations supporting its utility. As such, we hope that the idea described here along with the detailed discussion of the presented results might entice further experimentation to promote the scope of this approach. In our empirical example we have used participants enrolled in the UK Biobank, which due to its UK base, might partially overlap with the data used by the QResearch group. As such the observed performance estimates might be (slightly) optimistic. We expect however that this will impact all models to a similar degree and hence not impact our main findings .

## CONCLUSION

Here we describe a novel approach referred to as ”stacked transfer learning”, which combines multiple pre-trained risk prediction models to derive a locally optimized model. We argue that due to its data-efficient nature, stacked transfer learning can allow non-data rich organisations, such as moderately sized healthcare trusts, to tailor prediction models to their local setting and hence optimize healthcare decision-making. Further, we show that our proposed ”stacked transfer learning” trained on one disease outcome could be repurposed in a less data-intensive manner for related disease.

## Supporting information

Supplementary file to the manuscript

## Data Availability

1. Dataset obtained from the UK Biobank.
2. All data produced for this submission as figures, and tables are available in the main manuscript and the supplementary file.
3. All code scripts used for running experiments, and producing tables and figures will shortly be made available via GitLab.

https://www.ukbiobank.ac.uk/

https://gitlab.com/SchmidtAF/qscores_stacked

## Competing interests

AFS has received funding from New Amsterdam Pharma for unrelated work.

## Funding

Folkert Asselbergs and A. Floriaan Schmidt are supported by the UCL Hospital NIHR Biomedical Research Centre grant. A. Floriaan Schmidt is supported by British Heart Foundation (BHF) grant PG/22/10989 and the UCL BHF Research Accelerator AA/18/6/34223. This work received support from the Dutch Research Council (MyDigiTwin 628.011.213). This work was supported by the EXPANSE and EXPOSOME-NL projects. The EXPANSE project is funded by the European Union’s Horizon 2020 research and innovation programme under grand agreement No 874627. The EXPOSOME-NL project is funded through the Gravitation program of the Dutch Ministry of Education, Culture, and Science and the Netherlands Organization for Scientific Research (NWO grant number 024.004.017). Jasmine Gratton was supported by the BHF studentship FS/17/70/33482.

## Acknowledgement

This research has been conducted using the UK Biobank Resource under Application Numbers 12113 and 24711. The authors are grateful to UK Biobank participants. UK Biobank was established by the Wellcome Trust medical charity, Medical Research Council, Department of Health, Scottish Government, and the Northwest Regional Development Agency. It has also had funding from the Welsh Assembly Government and the British Heart Foundation. The authors also thank the administrators of the high performance computing cluster of UMC Utrecht.

## Notes

### Author Declarations

This research has been conducted using the UK Biobank Resource under Application Numbers 12113 and 24711.

