## Supplementary file to the manuscript for "Stacking multiple prediction models to optimise performance in local settings: exemplars in cardiometabolic disease"

### Appendix

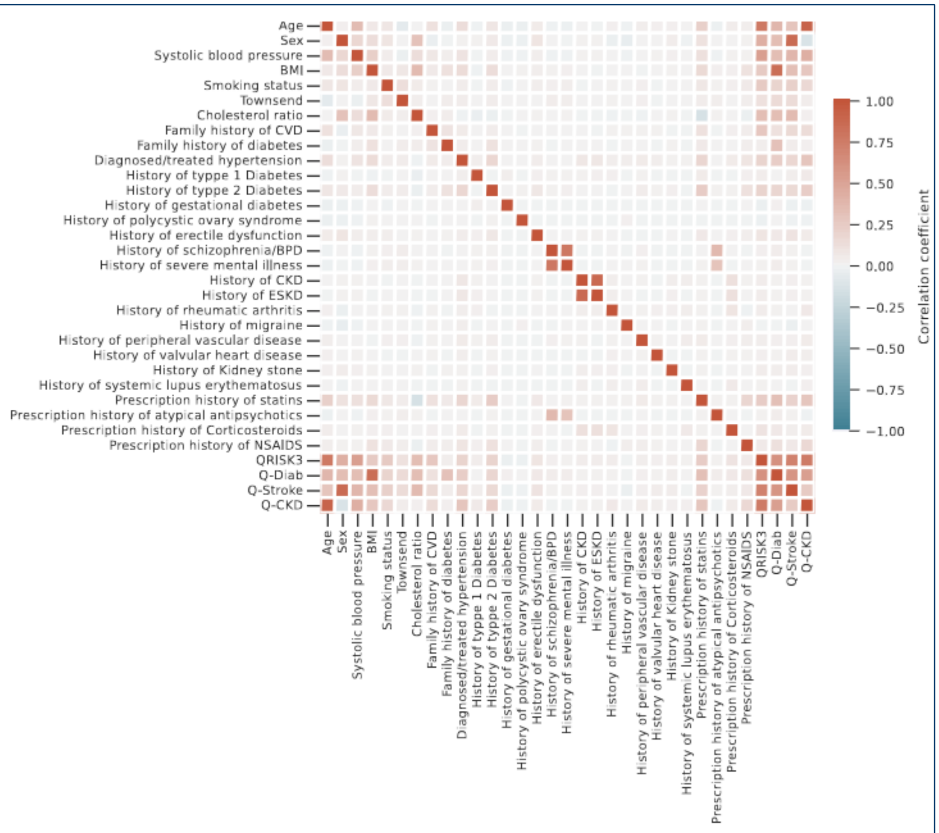


**Figure A1** The pairwise Spearman correlation coefficients between baseline characteristics and the considered Q-scores estimated in the 325,654 UKB participants without established CVD at baseline.

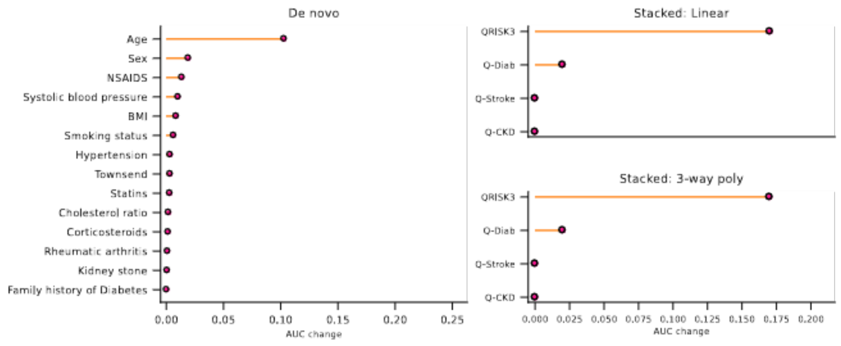


**Figure A2** Contribution of each of the predictors in the *de novo* model (left panel) and each of the Q-scores in the stacked models (right panel) for CVD incidence prediction. The stacked and *de novo* models were trained on an illustrative example dataset of 100 CVD cases (1384 samples in total). Please see Figure 2 and 3, and Table A3 for performance of the entire range of sample sizes.

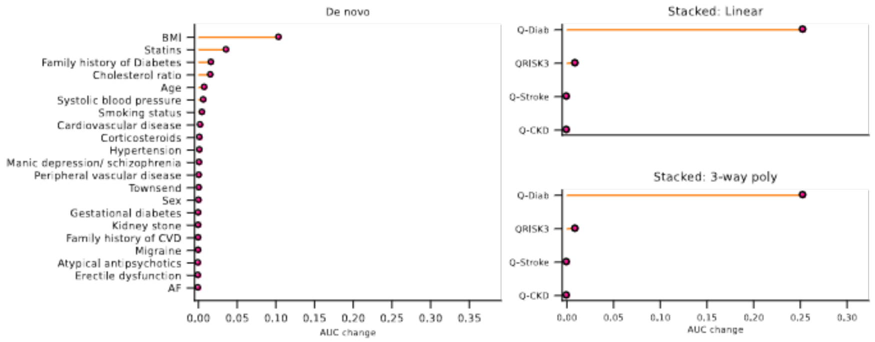


**Figure A3** Contribution of each of the predictors in the *de novo* model (left panel) and each of the Q-scores in the stacked models (right panel) for T2D incidence prediction. The stacked and *de novo* models were trained on an illustrative example dataset of 100 T2D cases (2583 samples in total). Please see Figure 5 and A5, and Table A7 for performance of the entire range of sample sizes.

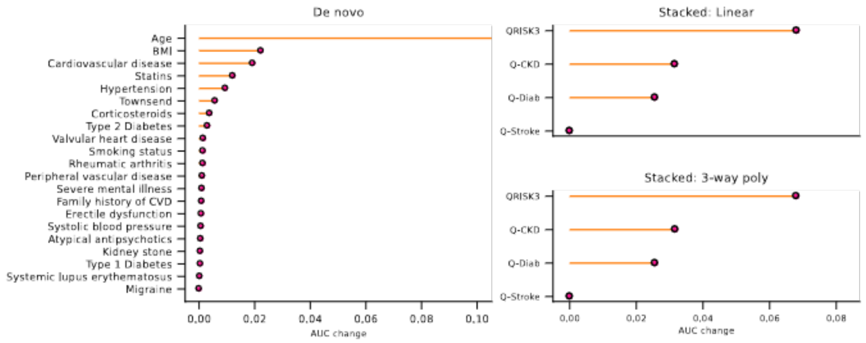


**Figure A4** Contribution of each of the predictors in the *de novo* model (left panel) and each of the Q-scores in the stacked models (right panel) for CKD incidence prediction. The stacked and *de novo* models were trained on an illustrative example dataset of 100 CKD cases (6698 samples in total). Please see Figure 5 and A6, and Table A8 for performance of the entire range of sample sizes.

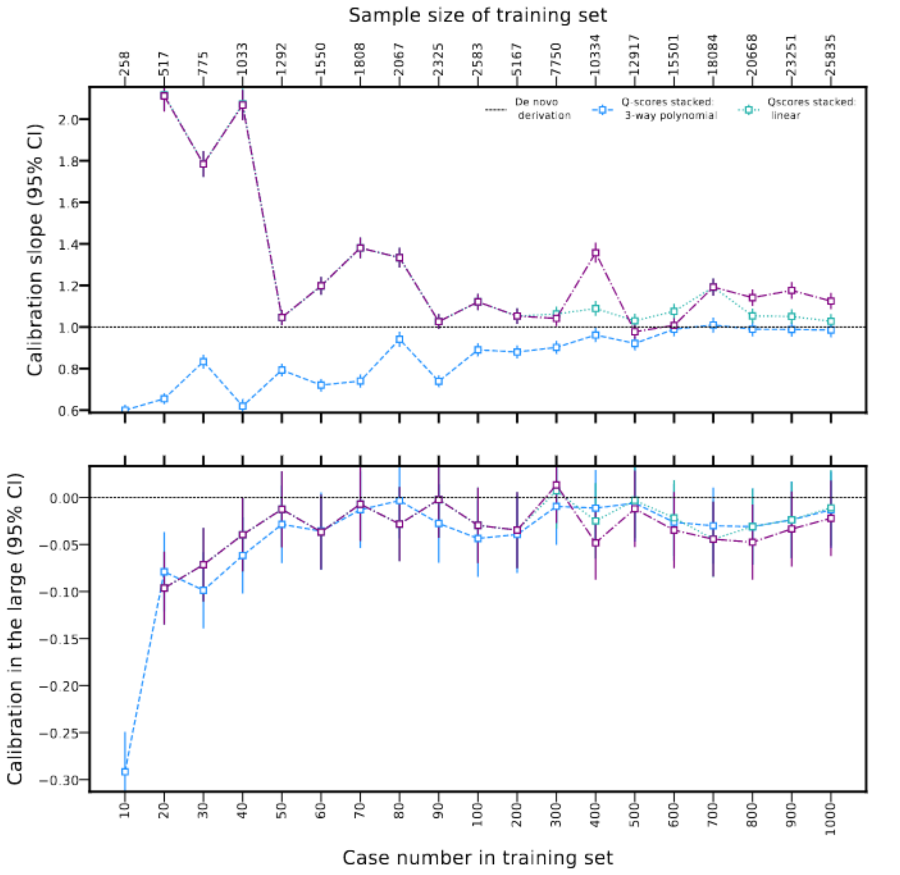


**Figure A5** Plots of the calibration estimates, illustrating the agreement between predicted T2D risk and observed T2D risks for the stacked Q-risk score models and the *de novo* model, across a range of training sample sizes and corresponding case numbers. Estimates were derived from an independent test set of 10,683 T2D cases and a total of 279,290 participants. The numerical data underpinning this figure are provided in Table A7.

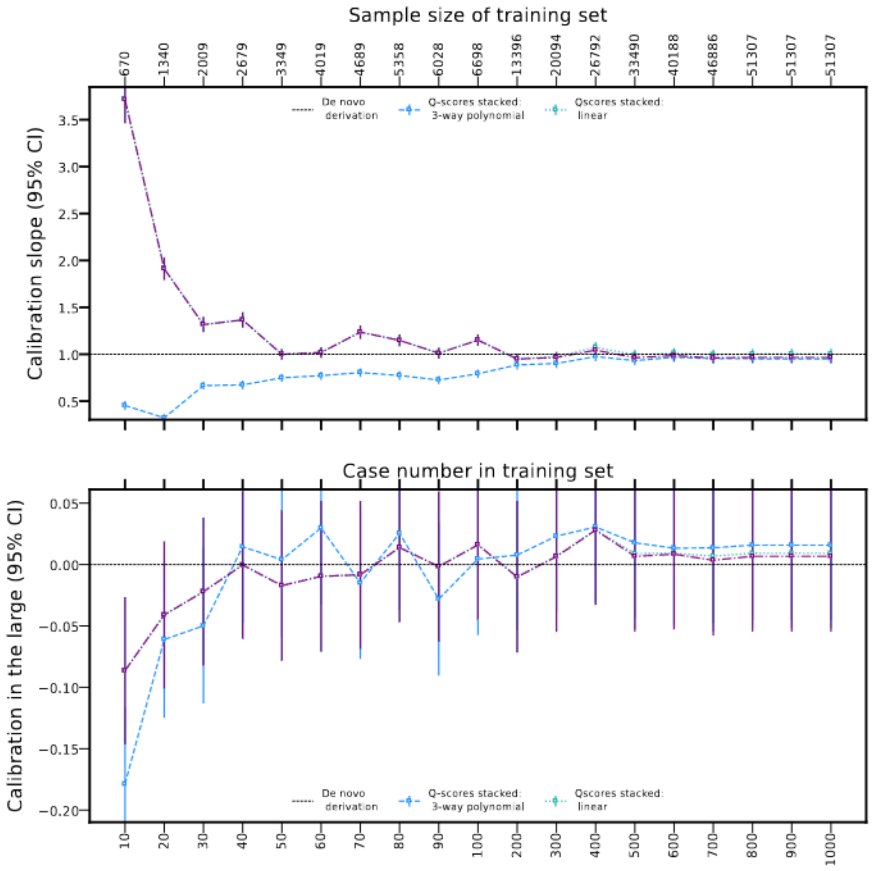


**Figure A6** Plots of the calibration estimates, illustrating the agreement between predicted CKD risk and observed CKD risks for the stacked Q-risk score models and the *de novo* model, across a range of training sample sizes and corresponding case numbers. Estimates were derived from an independent test set of 4390 CKD cases and a total of 289,594 participants. The numerical data underpinning this figure are provided in Table A8.

**Table A1** Variables used to calculate the Q-research scores and their corresponding UK biobank field number and ICD-10 codes.

| **Predictor variables** | **Field number** | **ICD-codes from field 41270** |
|---|---|---|
| sex | 31 | |
| age | 21003 | |
| ethnicity | 22006 | |
| Townsend deprivation index | 189 | |
| systolic BP | 93, 4080 | |
| BMI | 21001 | |
| smoking | 20116, 3456 | |
| type 1 diabetes | 41270 | E10 |
| type 2 diabetes | 41270 | E11 |
| treated hypertension | 20002, 41270, 20003 | I10 |
| total cholesterol:HDL cholesterol ratio | 30690, 30760 | |
| family history of CHD in a 1st degree relative $< 60$ yrs | 20107, 20110, 20111 | |
| rheumatoid arthritis | 41270 | M06.9, M05.0, M05.1, J99.0, M06.1, M06.4 |
| atrial fibrillation | 41270 | I48 |
| major chronic renal disease | 41270 | N04, N03.2, N11.1, Z99.2, Z94.0 |
| chronic kidney disease (stages 3, 4 or 5) | 41270 | N18.0, N18.3, N18.4, N18.5, N18.9 |
| migraine | 41270 | G43, G44.0 |
| corticosteroids | 20003 | |
| systemic lupus erythematosus | 41270 | M32.9, M32.1, I39 |
| atypical antipsychotics | 20003 | |
| severe mental illness | 41270 | F23, F28, F29, F20, F31 |
| erectile dysfunction diagnosis | 41270, 20003 | N52.9 |
| chronic kidney disease (stages 4 or 5) | 41270 | N18.0, N18.4, N18.5, N18.9 |
| congestive cardiac failure | 41270 | I50.0 |
| coronary heart disease | 41270 | G45, I20, I21, I22, I23, I24, I25 |
| valvular heart disease | 41270 | I34, I35, I05, I06, I08 |
| cardiovascular disease | 41270 | G45, I20, I21, I22, I23, I24, I25, I63, I64 |
| gestational diabetes | 41270 | O244, O249 |
| polycystic ovary syndrome | 41270 | E282 |
| statins | 20003 | |
| family history of diabetes | 20107 | |
| NSAIDs | 6154 | |
| peripheral vascular disease | 41270 | I73 |
| kidney stones | 41270 | N200, N202, N209 |

**Table A2** Variables needed (indicated by ✓) to calculate each of the Qrisk scores.

| **Preditor variable** | **QRISK3** | **Q-Stroke** | **Q-Diab** | **Q-CKD** |
|---|---|---|---|---|
| sex | ✓ | ✓ | ✓ | ✓ |
| age | ✓ | ✓ | ✓ | ✓ |
| ethnicity | ✓ | ✓ | ✓ | ✓ |
| Townsend deprivation index | ✓ | ✓ | ✓ | ✓ |
| systolic BP | ✓ | ✓ | ✓ | ✓ |
| BMI | ✓ | ✓ | ✓ | ✓ |
| smoking | ✓ | ✓ | ✓ | ✓ |
| type 1 diabetes | ✓ | ✓ | ✓ | ✓ |
| type 2 diabetes | ✓ | ✓ | ✓ | ✓ |
| treated hypertension | ✓ | ✓ | ✓ | ✓ |
| Chol ratio=total cholesterol:HDL cholesterol ratio | ✓ | ✓ | | |
| family history of CHD in a 1st degree relative $< 60$ yrs | ✓ | ✓ | | |
| rheumatoid arthritis | ✓ | ✓ | | ✓ |
| atrial fibrillation | ✓ | ✓ | | |
| major chronic renal disease | ✓ | | | |
| chronic kidney disease (stages 3, 4 or 5) | ✓ | | | |
| migraine | ✓ | | | |
| corticosteroids | ✓ | | ✓ | |
| systemic lupus erythematosus | ✓ | | | ✓ |
| atypical antipsychotics | ✓ | | ✓ | |
| severe mental illness | ✓ | | ✓ | |
| erectile dysfunction diagnosis | ✓ | | | |
| chronic kidney disease (stages 4 or 5) | | ✓ | | |
| congestive cardiac failure | | ✓ | | ✓ |
| coronary heart disease | | ✓ | | |
| valvular heart disease | | ✓ | | |
| cardiovascular disease | | | ✓ | ✓ |
| gestational diabetes | | | ✓ | |
| polycystic ovary syndrome | | | ✓ | |
| statins | | | ✓ | |
| family history of diabetes | | | ✓ | |
| NSAIDs | | | | ✓ |
| peripheral vascular disease | | | | ✓ |
| kidney stones | | | | ✓ |

**Table A3** Effect of decreasing training sample size on model discrimination for 10-year risk of CVD prediction, evaluated in a a hold-out tests set of $276,646$ participants (corresponding to Figure 2).

| $\mathbf{N_{cases}}$ $(\mathbf{N_{samples}})$ | **Model** | $\mathbf{N_{coefs}}$ | **AUC** $(\mathbf{95\%CI})$ | **Calib. slope** $(\mathbf{(95\%CI})$ | **Calib. in the large** $(\mathbf{95\%CI})$ |
|---|---|---|---|---|---|
| 10 (138) | Stacked: linear | 0 | 0.500 (0.496, 0.504) | NA | NA |
| | Stacked: 3-way poly | 0 | 0.500 (0.496, 0.504) | NA | NA |
| | De novo | 10 | 0.658 (0.655, 0.662) | 0.518 (0.505, 0.532) | -0.197 (-0.212, -0.181) |
| 20 (277) | Stacked: linear | 0 | 0.500 (0.496, 0.504) | NA | NA |
| | Stacked: 3-way poly | 0 | 0.500 (0.496, 0.504) | NA | NA |
| | De novo | 14 | 0.675 (0.671, 0.678) | 0.650 (0.634, 0.666) | -0.041 (-0.056, -0.026) |
| 30 (415) | Stacked: linear | 1 | 0.732 (0.728, 0.735) | 8.905 (8.738, 9.072) | -0.038 (-0.052, -0.024) |
| | Stacked: 3-way poly | 2 | 0.732 (0.728, 0.735) | 8.987 (8.818, 9.155) | -0.038 (-0.053, -0.024) |
| | De novo | 20 | 0.690 (0.687, 0.693) | 0.753 (0.737, 0.770) | 0.007 (-0.008, 0.022) |
| 40 (554) | Stacked: linear | 1 | 0.732 (0.728, 0.735) | 1.826 (1.791, 1.860) | -0.027 (-0.042, -0.013) |
| | Stacked: 3-way poly | 2 | 0.732 (0.728, 0.735) | 1.827 (1.793, 1.862) | -0.027 (-0.042, -0.013) |
| | De novo | 15 | 0.645 (0.641, 0.648) | 0.451 (0.438, 0.464) | 0.001 (-0.014, 0.016) |
| 50 (692) | Stacked: linear | 1 | 0.732 (0.728, 0.735) | 2.102 (2.063, 2.142) | -0.040 (-0.055, -0.026) |
| | Stacked: 3-way poly | 2 | 0.732 (0.728, 0.735) | 2.107 (2.068, 2.147) | -0.040 (-0.055, -0.026) |
| | De novo | 21 | 0.686 (0.683, 0.690) | 0.831 (0.812, 0.850) | -0.042 (-0.057, -0.027) |
| 60 (831) | Stacked: linear | 2 | 0.736 (0.733, 0.739) | 1.172 (1.150, 1.193) | -0.001 (-0.016, 0.014) |
| | Stacked: 3-way poly | 3 | 0.736 (0.733, 0.739) | 1.172 (1.151, 1.194) | -0.001 (-0.016, 0.014) |
| | De novo | 20 | 0.727 (0.724, 0.730) | 0.760 (0.746, 0.774) | 0.108 (0.093, 0.123) |
| 70 (969) | Stacked: linear | 2 | 0.733 (0.730, 0.736) | 1.960 (1.924, 1.997) | -0.002 (-0.017, 0.012) |
| | Stacked: 3-way poly | 3 | 0.733 (0.730, 0.736) | 1.961 (1.924, 1.997) | -0.002 (-0.017, 0.012) |
| | De novo | 23 | 0.702 (0.699, 0.705) | 0.552 (0.540, 0.564) | -0.032 (-0.048, -0.017) |
| 80 (1108) | Stacked: linear | 1 | 0.732 (0.728, 0.735) | 1.727 (1.695, 1.759) | 0.009 (-0.005, 0.024) |
| | Stacked: 3-way poly | 2 | 0.732 (0.728, 0.735) | 1.729 (1.697, 1.762) | 0.009 (-0.005, 0.024) |
| | De novo | 25 | 0.720 (0.717, 0.724) | 0.759 (0.743, 0.775) | 0.017 (0.002, 0.032) |
| 90 (1246) | Stacked: linear | 2 | 0.736 (0.732, 0.739) | 1.162 (1.141, 1.184) | -0.039 (-0.053, -0.024) |
| | Stacked: 3-way poly | 3 | 0.736 (0.732, 0.739) | 1.163 (1.141, 1.184) | -0.039 (-0.053, -0.024) |
| | De novo | 23 | 0.716 (0.713, 0.719) | 0.611 (0.597, 0.624) | -0.060 (-0.076, -0.045) |
| 100 (1384) | Stacked: linear | 2 | 0.735 (0.732, 0.738) | 1.122 (1.101, 1.142) | -0.015 (-0.029, 0.000) |
| | Stacked: 3-way poly | 3 | 0.735 (0.732, 0.738) | 1.123 (1.102, 1.143) | -0.015 (-0.029, 0.000) |
| | De novo | 24 | 0.710 (0.706, 0.713) | 0.679 (0.665, 0.693) | -0.011 (-0.026, 0.004) |
| 200 (2769) | Stacked: linear | 2 | 0.734 (0.731, 0.737) | 1.117 (1.096, 1.137) | 0.001 (-0.013, 0.016) |
| | Stacked: 3-way poly | 3 | 0.734 (0.731, 0.737) | 1.117 (1.096, 1.137) | 0.001 (-0.013, 0.016) |
| | De novo | 27 | 0.723 (0.720, 0.726) | 0.786 (0.771, 0.802) | -0.005 (-0.020, 0.010) |
| 300 (4153) | Stacked: linear | 3 | 0.736 (0.733, 0.739) | 1.043 (1.024, 1.062) | -0.006 (-0.021, 0.009) |
| | Stacked: 3-way poly | 4 | 0.736 (0.733, 0.739) | 1.043 (1.024, 1.062) | -0.006 (-0.021, 0.009) |
| | De novo | 25 | 0.733 (0.730, 0.736) | 0.827 (0.811, 0.842) | -0.010 (-0.025, 0.005) |
| 400 (5538) | Stacked: linear | 3 | 0.736 (0.733, 0.739) | 1.011 (0.993, 1.030) | -0.022 (-0.037, -0.007) |
| | Stacked: 3-way poly | 4 | 0.736 (0.733, 0.739) | 1.011 (0.992, 1.029) | -0.022 (-0.037, -0.007) |
| | De novo | 28 | 0.732 (0.729, 0.735) | 0.843 (0.828, 0.859) | -0.007 (-0.022, 0.008) |
| 500 (6922) | Stacked: linear | 3 | 0.737 (0.734, 0.740) | 1.003 (0.984, 1.021) | -0.015 (-0.030, -0.000) |
| | Stacked: 3-way poly | 5 | 0.737 (0.734, 0.740) | 1.004 (0.986, 1.023) | -0.015 (-0.030, -0.000) |
| | De novo | 28 | 0.741 (0.738, 0.744) | 0.901 (0.885, 0.917) | -0.004 (-0.019, 0.011) |
| 600 (8306) | Stacked: linear | 4 | 0.737 (0.734, 0.740) | 1.075 (1.055, 1.094) | 0.005 (-0.010, 0.020) |
| | Stacked: 3-way poly | 6 | 0.736 (0.733, 0.739) | 1.068 (1.049, 1.087) | -0.000 (-0.015, 0.015) |
| | De novo | 28 | 0.733 (0.730, 0.736) | 0.849 (0.833, 0.865) | 0.009 (-0.006, 0.024) |
| 700 (9691) | Stacked: linear | 4 | 0.736 (0.733, 0.739) | 1.044 (1.025, 1.063) | -0.002 (-0.017, 0.013) |
| | Stacked: 3-way poly | 6 | 0.737 (0.734, 0.740) | 1.088 (1.068, 1.107) | 0.000 (-0.014, 0.015) |
| | De novo | 28 | 0.739 (0.736, 0.742) | 0.905 (0.889, 0.922) | 0.001 (-0.014, 0.016) |
| 800 (11075) | Stacked: linear | 3 | 0.737 (0.734, 0.740) | 0.975 (0.957, 0.993) | -0.004 (-0.019, 0.011) |
| | Stacked: 3-way poly | 6 | 0.737 (0.734, 0.740) | 1.004 (0.986, 1.022) | -0.004 (-0.019, 0.011) |
| | De novo | 28 | 0.742 (0.739, 0.745) | 0.898 (0.882, 0.913) | -0.004 (-0.019, 0.011) |
| 900 (12460) | Stacked: linear | 4 | 0.737 (0.734, 0.740) | 1.042 (1.023, 1.061) | -0.008 (-0.023, 0.007) |
| | Stacked: 3-way poly | 7 | 0.737 (0.734, 0.740) | 1.076 (1.057, 1.095) | -0.009 (-0.024, 0.006) |
| | De novo | 29 | 0.741 (0.738, 0.744) | 0.913 (0.897, 0.929) | -0.010 (-0.025, 0.005) |
| 1000 (13844) | Stacked: linear | 3 | 0.736 (0.733, 0.739) | 0.999 (0.981, 1.017) | 0.001 (-0.014, 0.016) |
| | Stacked: 3-way poly | 5 | 0.737 (0.734, 0.740) | 1.019 (1.001, 1.037) | 0.001 (-0.013, 0.016) |
| | De novo | 29 | 0.745 (0.742, 0.748) | 0.940 (0.923, 0.956) | -0.001 (-0.016, 0.014) |

**Table A4** AUC, calibration slope and calibration intercept (calibration-in-the-large) estimates for the prediction of 10-years CVD risk by individual Q-scores and models trained on randomly selected 100 CVD cases (1384 samples in total) (corresponding to Figure 4).

| Model | AUC (95%CI) | Calib. slope (95%CI) | Calib. intercept (95%CI) |
|---|---|---|---|
| De novo | 0.710 (0.706 , 0.713) | 0.679 (0.665 , 0.693) | -0.011 (-0.026 , 0.004) |
| Stacked: 3-way poly | 0.735 (0.732 , 0.738) | 1.123 (1.102 , 1.143) | -0.015 (-0.029 , 0.000) |
| Stacked: Linear | 0.735 (0.732 , 0.738) | 1.122 (1.101 , 1.142) | -0.015 (-0.029 , 0.000) |
| Q-Diabetes | 0.674 (0.671 , 0.678) | 0.621 (0.606 , 0.636) | 0.135 (0.120 , 0.150) |
| Q-Stroke | 0.689 (0.686 , 0.693) | 0.318 (0.310 , 0.325) | 1.504 (1.488 , 1.519) |
| Q-CKD | 0.692 (0.688 , 0.695) | 0.662 (0.647 , 0.676) | 1.363 (1.348 , 1.378) |
| QRISK3 | 0.732 (0.729 , 0.735) | 1.023 (1.003 , 1.042) | -0.308 (-0.323 , -0.294) |

**Table A5** Baseline characteristics for participants in the training data for Type 2 Diabetes incidence prediction.

| Participant characteristics | Cases | Controls | Missing-ness % |
|---|---|---|---|
| Total sample size (n) | 12599 | 316190 | |
| Age (median [IQR]) | 61.0 (55.0 , 65.0) | 58.0 (50.0 , 63.0) | 0.0 |
| Sex (male) (%) | 7548.0 (59.91%) | 142601.0 (45.1%) | 0.0 |
| Systolic blood pressure in mmHg (median [IQR]) | 145.0 (133.0 , 159.0) | 138.0 (126.0 , 152.0) | 0.09 |
| HDL cholesterol in mmol/L (median [IQR]) | 1.16 (1.0 , 1.37) | 1.42 (1.2 , 1.7) | 12.73 |
| LDL cholesterol in mmol/L (median [IQR]) | 3.24 (2.61 , 3.96) | 3.57 (3.01 , 4.16) | 4.84 |
| Triglycerides in mmol/L (median [IQR]) | 2.13 (1.51 , 2.96) | 1.46 (1.04 , 2.1) | 4.74 |
| BMI in kg/m2 (median [IQR]) | 31.06 (28.0 , 34.93) | 26.47 (23.98 , 29.44) | 0.30 |
| Smoking status (%) | | | 0.04 |
| Non-smoker | 5431 ( 43.0 %) | 175384 (55.0%) | |
| Former smoker | 5393 ( 43.0 %) | 108910 ( 34.0 %) | |
| Light smoker ($< 10$ cigarettes/day) | 621 ( 5.0 %) | 8906 ( 3.0 %) | |
| Moderate smoker (10-19 cigarettes/day) | 489 ( 4.0 %) | 7671 ( 2.0 %) | |
| Heavy smoker ($> 20$ cigarettes/day) | 145 ( 1.0 %) | 4222 ( 1.0 %) | |
| Diagnosed/ treated hypertension | 1740.0 (13.81%) | 14526.0 (4.59%) | 0.0 |
| History of cardiovascular diseases | 1352.0 (10.73%) | 12158.0 (3.85%) | 0.0 |
| History of atrial fibrillation | 329.0 (2.61%) | 3809.0 (1.2%) | 0.0 |
| History of peripheral vascular disease | 110.0 (0.87%) | 836.0 (0.26%) | 0.0 |
| History of valvular heart disease | 110.0 (0.87%) | 1375.0 (0.43%) | 0.0 |
| History of chronic heart disease | 1051.0 (8.34%) | 8418.0 (2.66%) | 0.0 |
| History of congestive cardiac failure | 3.0 (0.02%) | 8.0 (0.0%) | 0.0 |
| History of rhematic arthritis | 86.0 (0.68%) | 1274.0 (0.4%) | 0.0 |
| History of Systemic lupus erythematosus | 6.0 (0.05%) | 171.0 (0.05%) | 0.0 |
| History of chronic kidney disease | 35.0 (0.28%) | 343.0 (0.11%) | 0.0 |
| History of end stage kidney disease | 35.0 (0.28%) | 343.0 (0.11%) | 0.0 |
| History of kidney stones | 107.0 (0.85%) | 1239.0 (0.39%) | 0.0 |
| History of migraine | 117.0 (0.93%) | 2557.0 (0.81%) | 0.0 |
| History of severe mental illness | 109.0 (0.87%) | 1034.0 (0.33%) | 0.0 |
| Prescription history of Statins | 4266.0 (33.86%) | 34884.0 (11.03%) | 0.0 |
| Prescription history of atypical antipsychotics | 99.0 (0.79%) | 759.0 (0.24%) | 0.0 |
| Prescription history of corticosteroids | 341.0 (2.71%) | 4770.0 (1.51%) | 0.0 |
| Prescription history of NSAID | 5097.0 (40.46%) | 80882.0 (25.58%) | 0.0 |

**Table A6** Baseline characteristics of participants in the training data for chronic kidney disease incidence prediction.

| **Participant characteristics** | **Cases** | **Controls** | **Missing-ness (%)** |
|---|---|---|---|
| Total sample size (n) | 5156 | 335745 | |
| Age (median [IQR]) | 64.0 (60.0 , 67.0) | 58.0 (51.0 , 63.0) | 0.0 |
| Sex (male) (%) | 2764.0 (53.61%) | 154712.0 (46.08%) | 0.0 |
| Systolic blood pressure in mmHg (median [IQR]) | 144.0 (131.0 , 158.0) | 139.0 (126.0 , 152.0) | 0.09 |
| HDL cholesterol in mmol/L (median [IQR]) | 1.23 (1.03 , 1.49) | 1.41 (1.18 , 1.68) | 12.76 |
| LDL cholesterol in mmol/L (median [IQR]) | 3.06 (2.48 , 3.82) | 3.54 (2.97 , 4.14) | 4.85 |
| Triglycerides in mmol/L (median [IQR]) | 1.81 (1.27 , 2.57) | 1.49 (1.05 , 2.15) | 4.75 |
| BMI in kg/m2 (median [IQR]) | 29.24 (26.08 , 33.04) | 26.68 (24.11 , 29.79) | 0.32 |
| Smoking status (%) | | | 0.04 |
| Non-smoker | 2354 ( 46.0 %) | 183822 (55.0%) | |
| Former smoker | 2199 ( 43.0 %) | 117459 ( 35.0 %) | |
| Light smoker ($< 10$ cigarettes/day) | 169 ( 3.0 %) | 9602 ( 3.0 %) | |
| Moderate smoker (10-19 cigarettes/day) | 158 ( 3.0 %) | 8550 ( 3.00 %) | |
| Heavy smoker ($> 20$ cigarettes/day) | 65 ( 1.0 %) | 4408 ( 1.0 %) | |
| Diagnosed/ treated hypertension | 1418.0 (27.5%) | 18220.0 (5.43%) | 0.0 |
| History of Type 1 Diabetes | 21.0 (0.41%) | 456.0 (0.14%) | 0.0 |
| History of Type 2 Diabetes | 697.0 (13.52%) | 7946.0 (2.37%) | 0.0 |
| History of cardiovascular diseases | 1107.0 (21.47%) | 14559.0 (4.34%) | 0.0 |
| History of atrial fibrillation | 349.0 (6.77%) | 4278.0 (1.27%) | 0.0 |
| History of peripheral vascular disease | 115.0 (2.23%) | 1040.0 (0.31%) | 0.0 |
| History of valvular heart disease | 142.0 (2.75%) | 1502.0 (0.45%) | 0.0 |
| History of chronic heart disease | 837.0 (16.23%) | 10424.0 (3.1%) | 0.0 |
| History of congestive cardiac failure | 3.0 (0.06%) | 12.0 (0.0%) | 0.0 |
| History of rhematic arthritis | 100.0 (1.94%) | 1391.0 (0.41%) | 0.0 |
| History of Systemic lupus erythematosus | 12.0 (0.23%) | 168.0 (0.05%) | 0.0 |
| History of kidney stones | 76.0 (1.47%) | 1430.0 (0.43%) | 0.0 |
| History of migraine | 54.0 (1.05%) | 2761.0 (0.82%) | 0.0 |
| History of severe mental illness | 60.0 (1.16%) | 1189.0 (0.35%) | 0.0 |
| Prescription history of Statins | 1854.0 (35.96%) | 43737.0 (13.03%) | 0.0 |
| Prescription history of atypical antipsychotics | 55.0 (1.07%) | 863.0 (0.26%) | 0.0 |
| Prescription history of corticosteroids | 253.0 (4.91%) | 5078.0 (1.51%) | 0.0 |
| Prescription history of NSAID | 2212.0 (42.9%) | 90477.0 (26.95%) | 0.0 |

**Table A7** Effect of decreasing training sample size on model discrimination for 10-year risk of T2D prediction, evaluated in a hold-out tests set of $279,290$ participants (corresponding to Figure 5).

| $N_{cases}$ ($N_{samples}$) | Model | $N_{coefs}$ | AUC ($95\%CI$) | Calib. slope (($95\%CI$) | Calib. in the large ($95\%CI$) |
|---|---|---|---|---|---|
| 10 (258) | stacked: simple linear | 4 | 0.792 (0.788, 0.795) | 0.631 (0.619, 0.642) | -0.149 (-0.171, -0.128) |
| | Stacked: linear | 0 | 0.500 (0.494, 0.506) | NA | NA |
| | Stacked: 3-way poly | 0 | 0.500 (0.494, 0.506) | NA | NA |
| | De novo | 17 | 0.767 (0.763, 0.770) | 0.600 (0.588, 0.612) | -0.292 (-0.313, -0.271) |
| 20 (517) | stacked: simple linear | 4 | 0.807 (0.804, 0.810) | 0.796 (0.782, 0.810) | -0.148 (-0.169, -0.127) |
| | Stacked: linear | 1 | 0.815 (0.812, 0.818) | 2.112 (2.074, 2.149) | -0.096 (-0.116, -0.077) |
| | Stacked: 3-way poly | 2 | 0.815 (0.812, 0.818) | 2.117 (2.080, 2.155) | -0.096 (-0.116, -0.077) |
| | De novo | 18 | 0.782 (0.779, 0.786) | 0.655 (0.642, 0.668) | -0.079 (-0.100, -0.058) |
| 30 (775) | stacked: simple linear | 4 | 0.798 (0.795, 0.802) | 0.913 (0.897, 0.930) | -0.072 (-0.092, -0.052) |
| | Stacked: linear | 2 | 0.812 (0.809, 0.815) | 1.784 (1.753, 1.816) | -0.072 (-0.091, -0.052) |
| | Stacked: 3-way poly | 3 | 0.812 (0.809, 0.815) | 1.784 (1.753, 1.816) | -0.071 (-0.091, -0.052) |
| | De novo | 20 | 0.778 (0.775, 0.782) | 0.834 (0.817, 0.850) | -0.099 (-0.119, -0.078) |
| 40 (1033) | stacked: simple linear | 4 | 0.756 (0.752, 0.759) | 1.006 (0.986, 1.027) | -0.033 (-0.053, -0.013) |
| | Stacked: linear | 1 | 0.815 (0.812, 0.818) | 2.068 (2.031, 2.104) | -0.040 (-0.059, -0.020) |
| | Stacked: 3-way poly | 2 | 0.815 (0.812, 0.818) | 2.074 (2.037, 2.110) | -0.040 (-0.059, -0.020) |
| | De novo | 21 | 0.732 (0.728, 0.736) | 0.620 (0.603, 0.636) | -0.062 (-0.082, -0.041) |
| 50 (1292) | stacked: simple linear | 4 | 0.817 (0.814, 0.820) | 0.856 (0.841, 0.871) | 0.021 (0.000, 0.042) |
| | Stacked: linear | 1 | 0.815 (0.812, 0.818) | 1.046 (1.028, 1.065) | -0.013 (-0.033, 0.008) |
| | Stacked: 3-way poly | 2 | 0.815 (0.812, 0.818) | 1.046 (1.027, 1.064) | -0.013 (-0.033, 0.008) |
| | De novo | 23 | 0.798 (0.795, 0.801) | 0.793 (0.778, 0.808) | -0.029 (-0.049, -0.008) |
| 60 (1550) | stacked: simple linear | 4 | 0.797 (0.793, 0.800) | 0.906 (0.890, 0.923) | -0.020 (-0.040, 0.001) |
| | Stacked: linear | 2 | 0.805 (0.801, 0.808) | 1.198 (1.177, 1.219) | -0.037 (-0.057, -0.017) |
| | Stacked: 3-way poly | 3 | 0.805 (0.801, 0.808) | 1.198 (1.176, 1.219) | -0.037 (-0.057, -0.017) |
| | De novo | 28 | 0.774 (0.770, 0.777) | 0.721 (0.706, 0.735) | -0.036 (-0.056, -0.015) |
| 70 (1808) | stacked: simple linear | 4 | 0.797 (0.794, 0.800) | 1.072 (1.053, 1.092) | 0.013 (-0.007, 0.033) |
| | Stacked: linear | 2 | 0.800 (0.797, 0.803) | 1.381 (1.356, 1.405) | -0.007 (-0.027, 0.013) |
| | Stacked: 3-way poly | 3 | 0.800 (0.797, 0.803) | 1.380 (1.355, 1.405) | -0.007 (-0.027, 0.013) |
| | De novo | 27 | 0.773 (0.769, 0.776) | 0.740 (0.724, 0.756) | -0.013 (-0.033, 0.007) |
| 80 (2067) | stacked: simple linear | 4 | 0.810 (0.807, 0.813) | 1.114 (1.094, 1.133) | -0.017 (-0.037, 0.003) |
| | Stacked: linear | 1 | 0.815 (0.812, 0.818) | 1.334 (1.310, 1.357) | -0.028 (-0.048, -0.008) |
| | Stacked: 3-way poly | 2 | 0.815 (0.812, 0.818) | 1.334 (1.310, 1.358) | -0.028 (-0.048, -0.008) |
| | De novo | 26 | 0.785 (0.782, 0.789) | 0.941 (0.923, 0.959) | -0.003 (-0.023, 0.017) |
| 90 (2325) | stacked: simple linear | 4 | 0.817 (0.814, 0.820) | 0.932 (0.916, 0.949) | 0.013 (-0.007, 0.034) |
| | Stacked: linear | 1 | 0.815 (0.812, 0.818) | 1.026 (1.008, 1.044) | -0.002 (-0.023, 0.018) |
| | Stacked: 3-way poly | 2 | 0.815 (0.812, 0.818) | 1.027 (1.008, 1.045) | -0.002 (-0.023, 0.018) |
| | De novo | 25 | 0.797 (0.793, 0.800) | 0.739 (0.725, 0.753) | -0.028 (-0.048, -0.007) |
| 100 (2583) | stacked: simple linear | 4 | 0.809 (0.806, 0.812) | 0.980 (0.963, 0.998) | -0.013 (-0.033, 0.007) |
| | Stacked: linear | 2 | 0.816 (0.813, 0.819) | 1.121 (1.101, 1.141) | -0.030 (-0.050, -0.010) |
| | Stacked: 3-way poly | 3 | 0.816 (0.813, 0.819) | 1.121 (1.101, 1.141) | -0.030 (-0.050, -0.010) |
| | De novo | 25 | 0.813 (0.810, 0.816) | 0.890 (0.875, 0.906) | -0.044 (-0.064, -0.023) |
| 200 (5167) | stacked: simple linear | 4 | 0.812 (0.809, 0.815) | 0.972 (0.955, 0.990) | -0.033 (-0.054, -0.013) |
| | Stacked: linear | 2 | 0.817 (0.813, 0.820) | 1.053 (1.034, 1.071) | -0.035 (-0.055, -0.015) |
| | Stacked: 3-way poly | 3 | 0.817 (0.813, 0.820) | 1.052 (1.034, 1.071) | -0.035 (-0.055, -0.015) |
| | De novo | 29 | 0.810 (0.807, 0.814) | 0.880 (0.864, 0.895) | -0.039 (-0.060, -0.019) |
| 300 (7750) | stacked: simple linear | 4 | 0.810 (0.806, 0.813) | 0.994 (0.977, 1.012) | 0.016 (-0.005, 0.036) |
| | Stacked: linear | 2 | 0.810 (0.807, 0.814) | 1.041 (1.023, 1.060) | 0.013 (-0.007, 0.033) |
| | Stacked: 3-way poly | 4 | 0.815 (0.812, 0.818) | 1.063 (1.045, 1.080) | 0.007 (-0.013, 0.027) |
| | De novo | 29 | 0.818 (0.815, 0.821) | 0.902 (0.886, 0.918) | -0.009 (-0.030, 0.011) |
| 400 (10334) | stacked: simple linear | 4 | 0.814 (0.810, 0.817) | 0.999 (0.981, 1.017) | -0.028 (-0.048, -0.007) |
| | Stacked: linear | 1 | 0.815 (0.812, 0.818) | 1.358 (1.333, 1.382) | -0.048 (-0.068, -0.028) |
| | Stacked: 3-way poly | 4 | 0.816 (0.813, 0.819) | 1.089 (1.071, 1.107) | -0.025 (-0.045, -0.005) |
| | De novo | 31 | 0.816 (0.813, 0.819) | 0.961 (0.945, 0.978) | -0.011 (-0.032, 0.009) |
| 500 (12917) | stacked: simple linear | 4 | 0.815 (0.812, 0.818) | 0.956 (0.939, 0.972) | -0.008 (-0.029, 0.012) |
| | Stacked: linear | 3 | 0.815 (0.812, 0.818) | 0.977 (0.960, 0.994) | -0.012 (-0.032, 0.008) |
| | Stacked: 3-way poly | 5 | 0.817 (0.813, 0.820) | 1.030 (1.013, 1.047) | -0.003 (-0.023, 0.017) |
| | De novo | 29 | 0.819 (0.816, 0.822) | 0.921 (0.905, 0.937) | -0.006 (-0.026, 0.014) |
| 600 (15501) | stacked: simple linear | 4 | 0.813 (0.810, 0.816) | 0.983 (0.966, 1.001) | -0.031 (-0.052, -0.011) |
| | Stacked: linear | 3 | 0.814 (0.811, 0.817) | 1.008 (0.990, 1.025) | -0.035 (-0.055, -0.014) |
| | Stacked: 3-way poly | 5 | 0.817 (0.813, 0.820) | 1.075 (1.057, 1.093) | -0.022 (-0.042, -0.002) |
| | De novo | 30 | 0.823 (0.820, 0.826) | 0.989 (0.972, 1.006) | -0.026 (-0.047, -0.006) |
| 700 (18084) | stacked: simple linear | 4 | 0.813 (0.810, 0.816) | 0.995 (0.977, 1.012) | -0.034 (-0.054, -0.013) |
| | Stacked: linear | 2 | 0.816 (0.813, 0.819) | 1.192 (1.171, 1.213) | -0.044 (-0.064, -0.024) |
| | Stacked: 3-way poly | 3 | 0.816 (0.813, 0.819) | 1.192 (1.171, 1.213) | -0.044 (-0.064, -0.024) |
| | De novo | 30 | 0.822 (0.819, 0.825) | 1.010 (0.992, 1.027) | -0.030 (-0.050, -0.010) |
| 800 (20668) | stacked: simple linear | 4 | 0.813 (0.810, 0.816) | 0.974 (0.957, 0.991) | -0.041 (-0.061, -0.021) |
| | Stacked: linear | 2 | 0.816 (0.813, 0.819) | 1.141 (1.121, 1.161) | -0.048 (-0.068, -0.028) |
| | Stacked: 3-way poly | 5 | 0.817 (0.814, 0.820) | 1.053 (1.036, 1.071) | -0.031 (-0.051, -0.011) |
| | De novo | 30 | 0.823 (0.820, 0.826) | 0.989 (0.972, 1.006) | -0.031 (-0.051, -0.011) |
| 900 (23251) | stacked: simple linear | 4 | 0.812 (0.809, 0.815) | 1.014 (0.996, 1.032) | -0.022 (-0.042, -0.001) |
| | Stacked: linear | 2 | 0.815 (0.812, 0.818) | 1.176 (1.155, 1.197) | -0.033 (-0.053, -0.013) |
| | Stacked: 3-way poly | 6 | 0.817 (0.814, 0.820) | 1.051 (1.033, 1.068) | -0.024 (-0.044, -0.004) |
| | De novo | 31 | 0.823 (0.820, 0.826) | 0.988 (0.971, 1.005) | -0.024 (-0.044, -0.003) |
| 1000 (25835) | stacked: simple linear | 4 | 0.812 (0.809, 0.815) | 0.987 (0.970, 1.005) | -0.013 (-0.033, 0.008) |
| | Stacked: linear | 2 | 0.816 (0.813, 0.819) | 1.125 (1.105, 1.145) | -0.022 (-0.042, -0.002) |
| | Stacked: 3-way poly | 6 | 0.817 (0.814, 0.820) | 1.028 (1.010, 1.045) | -0.011 (-0.031, 0.009) |
| | De novo | 30 | 0.822 (0.819, 0.825) | 0.985 (0.968, 1.002) | -0.013 (-0.033, 0.007) |

**Table A8** Effect of decreasing training sample size on model discrimination for 5-year risk of CKD prediction, evaluated in a hold-out tests set of $289,594$ participants (corresponding to Figure 5).

| $\mathbf{N_{cases}}$ $\mathbf{(N_{samples})}$ | Model | $\mathbf{N_{coefs}}$ | AUC $\mathbf{(95\%CI)}$ | Calib. slope $\mathbf{((95\%CI)}$ | Calib. in the large $\mathbf{(95\%CI)}$ |
|---|---|---|---|---|---|
| 10 (670) | stacked: simple linear | 4 | 0.679 (0.672, 0.686) | 0.588 (0.562, 0.613) | -0.072 (-0.103, -0.041) |
| | Stacked: linear | 1 | 0.732 (0.726, 0.738) | 3.715 (3.589, 3.841) | -0.086 (-0.116, -0.057) |
| | Stacked: 3-way poly | 2 | 0.732 (0.726, 0.738) | 3.723 (3.596, 3.849) | -0.086 (-0.116, -0.057) |
| | De novo | 13 | 0.621 (0.613, 0.628) | 0.453 (0.430, 0.477) | -0.179 (-0.210, -0.148) |
| 20 (1340) | stacked: simple linear | 4 | 0.768 (0.762, 0.773) | 0.894 (0.869, 0.920) | 0.001 (-0.030, 0.031) |
| | Stacked: linear | 1 | 0.747 (0.741, 0.753) | 1.912 (1.852, 1.971) | -0.041 (-0.071, -0.011) |
| | Stacked: 3-way poly | 2 | 0.747 (0.741, 0.753) | 1.914 (1.854, 1.974) | -0.041 (-0.071, -0.011) |
| | De novo | 21 | 0.628 (0.621, 0.636) | 0.322 (0.302, 0.341) | -0.061 (-0.093, -0.029) |
| 30 (2009) | stacked: simple linear | 4 | 0.779 (0.774, 0.784) | 0.786 (0.765, 0.808) | -0.052 (-0.083, -0.021) |
| | Stacked: linear | 2 | 0.757 (0.751, 0.762) | 1.316 (1.277, 1.355) | -0.022 (-0.052, 0.008) |
| | Stacked: 3-way poly | 3 | 0.757 (0.751, 0.762) | 1.316 (1.277, 1.355) | -0.022 (-0.052, 0.008) |
| | De novo | 20 | 0.764 (0.758, 0.769) | 0.665 (0.647, 0.684) | -0.050 (-0.082, -0.018) |
| 40 (2679) | stacked: simple linear | 4 | 0.774 (0.768, 0.779) | 1.015 (0.987, 1.044) | 0.037 (0.006, 0.067) |
| | Stacked: linear | 2 | 0.762 (0.756, 0.767) | 1.366 (1.326, 1.406) | -0.000 (-0.030, 0.030) |
| | Stacked: 3-way poly | 3 | 0.762 (0.756, 0.767) | 1.367 (1.327, 1.407) | -0.000 (-0.030, 0.030) |
| | De novo | 28 | 0.728 (0.721, 0.734) | 0.674 (0.650, 0.697) | 0.014 (-0.016, 0.045) |
| 50 (3349) | stacked: simple linear | 4 | 0.778 (0.773, 0.783) | 0.760 (0.739, 0.781) | -0.017 (-0.048, 0.015) |
| | Stacked: linear | 3 | 0.780 (0.775, 0.785) | 0.999 (0.972, 1.026) | -0.017 (-0.048, 0.013) |
| | Stacked: 3-way poly | 4 | 0.780 (0.775, 0.785) | 0.999 (0.972, 1.026) | -0.017 (-0.048, 0.013) |
| | De novo | 26 | 0.788 (0.783, 0.794) | 0.749 (0.728, 0.769) | 0.004 (-0.027, 0.035) |
| 60 (4019) | stacked: simple linear | 4 | 0.791 (0.785, 0.796) | 0.813 (0.791, 0.836) | 0.011 (-0.020, 0.042) |
| | Stacked: linear | 3 | 0.788 (0.782, 0.793) | 1.019 (0.991, 1.046) | -0.010 (-0.040, 0.021) |
| | Stacked: 3-way poly | 4 | 0.788 (0.782, 0.793) | 1.019 (0.991, 1.046) | -0.010 (-0.040, 0.021) |
| | De novo | 28 | 0.775 (0.770, 0.780) | 0.772 (0.751, 0.794) | 0.030 (-0.001, 0.061) |
| 70 (4689) | stacked: simple linear | 4 | 0.778 (0.772, 0.783) | 1.015 (0.988, 1.043) | -0.011 (-0.041, 0.020) |
| | Stacked: linear | 2 | 0.764 (0.758, 0.769) | 1.236 (1.200, 1.272) | -0.008 (-0.038, 0.022) |
| | Stacked: 3-way poly | 3 | 0.764 (0.758, 0.769) | 1.236 (1.201, 1.272) | -0.008 (-0.038, 0.022) |
| | De novo | 24 | 0.776 (0.770, 0.781) | 0.804 (0.782, 0.827) | -0.015 (-0.046, 0.016) |
| 80 (5358) | stacked: simple linear | 4 | 0.787 (0.781, 0.792) | 0.958 (0.932, 0.984) | 0.033 (0.002, 0.064) |
| | Stacked: linear | 3 | 0.784 (0.779, 0.789) | 1.148 (1.118, 1.179) | 0.014 (-0.016, 0.044) |
| | Stacked: 3-way poly | 4 | 0.784 (0.779, 0.789) | 1.148 (1.118, 1.179) | 0.014 (-0.016, 0.044) |
| | De novo | 27 | 0.763 (0.758, 0.769) | 0.774 (0.752, 0.796) | 0.026 (-0.005, 0.056) |
| 90 (6028) | stacked: simple linear | 4 | 0.780 (0.775, 0.785) | 0.881 (0.857, 0.905) | 0.007 (-0.024, 0.038) |
| | Stacked: linear | 3 | 0.775 (0.770, 0.781) | 1.012 (0.984, 1.040) | -0.002 (-0.032, 0.029) |
| | Stacked: 3-way poly | 4 | 0.775 (0.770, 0.781) | 1.011 (0.984, 1.039) | -0.002 (-0.032, 0.029) |
| | De novo | 24 | 0.777 (0.772, 0.783) | 0.725 (0.705, 0.746) | -0.028 (-0.059, 0.003) |
| 100 (6698) | stacked: simple linear | 4 | 0.782 (0.777, 0.787) | 0.997 (0.970, 1.024) | 0.029 (-0.002, 0.059) |
| | Stacked: linear | 3 | 0.784 (0.778, 0.789) | 1.151 (1.120, 1.181) | 0.016 (-0.014, 0.046) |
| | Stacked: 3-way poly | 4 | 0.784 (0.778, 0.789) | 1.151 (1.120, 1.182) | 0.016 (-0.014, 0.046) |
| | De novo | 27 | 0.788 (0.783, 0.793) | 0.792 (0.770, 0.813) | 0.004 (-0.026, 0.035) |
| 200 (13396) | stacked: simple linear | 4 | 0.783 (0.777, 0.788) | 0.880 (0.856, 0.904) | -0.007 (-0.038, 0.024) |
| | Stacked: linear | 4 | 0.782 (0.777, 0.787) | 0.950 (0.924, 0.976) | -0.010 (-0.041, 0.021) |
| | Stacked: 3-way poly | 5 | 0.782 (0.777, 0.787) | 0.950 (0.924, 0.976) | -0.010 (-0.041, 0.021) |
| | De novo | 28 | 0.798 (0.793, 0.803) | 0.886 (0.863, 0.909) | 0.008 (-0.023, 0.038) |
| 300 (20094) | stacked: simple linear | 4 | 0.783 (0.778, 0.788) | 0.926 (0.901, 0.951) | 0.009 (-0.022, 0.040) |
| | Stacked: linear | 3 | 0.782 (0.777, 0.787) | 0.965 (0.939, 0.991) | 0.007 (-0.024, 0.037) |
| | Stacked: 3-way poly | 7 | 0.782 (0.777, 0.787) | 0.975 (0.949, 1.001) | 0.007 (-0.024, 0.037) |
| | De novo | 31 | 0.792 (0.787, 0.797) | 0.901 (0.878, 0.925) | 0.023 (-0.008, 0.054) |
| 400 (26792) | stacked: simple linear | 4 | 0.784 (0.779, 0.789) | 1.014 (0.986, 1.041) | 0.032 (0.001, 0.063) |
| | Stacked: linear | 3 | 0.784 (0.778, 0.789) | 1.044 (1.016, 1.072) | 0.028 (-0.002, 0.058) |
| | Stacked: 3-way poly | 7 | 0.783 (0.777, 0.788) | 1.074 (1.046, 1.102) | 0.028 (-0.002, 0.058) |
| | De novo | 31 | 0.804 (0.799, 0.809) | 0.975 (0.950, 1.000) | 0.031 (0.000, 0.061) |
| 500 (33490) | stacked: simple linear | 4 | 0.784 (0.779, 0.789) | 0.940 (0.914, 0.965) | 0.008 (-0.023, 0.039) |
| | Stacked: linear | 3 | 0.784 (0.778, 0.789) | 0.963 (0.937, 0.989) | 0.007 (-0.024, 0.037) |
| | Stacked: 3-way poly | 7 | 0.783 (0.777, 0.788) | 0.995 (0.969, 1.021) | 0.009 (-0.022, 0.039) |
| | De novo | 31 | 0.801 (0.796, 0.806) | 0.932 (0.908, 0.956) | 0.018 (-0.013, 0.048) |
| 600 (40188) | stacked: simple linear | 4 | 0.783 (0.778, 0.789) | 0.967 (0.941, 0.993) | 0.010 (-0.020, 0.041) |
| | Stacked: linear | 3 | 0.783 (0.777, 0.788) | 0.985 (0.959, 1.012) | 0.008 (-0.022, 0.039) |
| | Stacked: 3-way poly | 7 | 0.782 (0.777, 0.787) | 1.012 (0.985, 1.038) | 0.009 (-0.021, 0.040) |
| | De novo | 31 | 0.803 (0.798, 0.808) | 0.967 (0.943, 0.992) | 0.013 (-0.017, 0.044) |
| 700 (46886) | stacked: simple linear | 4 | 0.785 (0.780, 0.790) | 0.944 (0.919, 0.969) | 0.005 (-0.026, 0.035) |
| | Stacked: linear | 3 | 0.785 (0.779, 0.790) | 0.960 (0.935, 0.986) | 0.004 (-0.027, 0.034) |
| | Stacked: 3-way poly | 8 | 0.784 (0.778, 0.789) | 0.996 (0.970, 1.022) | 0.007 (-0.024, 0.037) |
| | De novo | 31 | 0.804 (0.799, 0.809) | 0.952 (0.928, 0.977) | 0.014 (-0.017, 0.044) |
| 800 (51307) | stacked: simple linear | 4 | 0.784 (0.779, 0.789) | 0.952 (0.927, 0.978) | 0.008 (-0.023, 0.039) |
| | Stacked: linear | 3 | 0.784 (0.778, 0.789) | 0.967 (0.941, 0.993) | 0.007 (-0.024, 0.037) |
| | Stacked: 3-way poly | 8 | 0.784 (0.779, 0.789) | 1.004 (0.978, 1.030) | 0.009 (-0.021, 0.040) |
| | De novo | 31 | 0.803 (0.799, 0.808) | 0.952 (0.927, 0.976) | 0.016 (-0.015, 0.046) |
| 900 (51307) | stacked: simple linear | 4 | 0.784 (0.779, 0.789) | 0.952 (0.927, 0.978) | 0.008 (-0.023, 0.039) |
| | Stacked: linear | 3 | 0.784 (0.778, 0.789) | 0.967 (0.941, 0.993) | 0.007 (-0.024, 0.037) |
| | Stacked: 3-way poly | 8 | 0.784 (0.779, 0.789) | 1.004 (0.978, 1.030) | 0.009 (-0.021, 0.040) |
| | De novo | 31 | 0.803 (0.799, 0.808) | 0.952 (0.927, 0.976) | 0.016 (-0.015, 0.046) |
| 1000 (51307) | stacked: simple linear | 4 | 0.784 (0.779, 0.789) | 0.952 (0.927, 0.978) | 0.008 (-0.023, 0.039) |
| | Stacked: linear | 3 | 0.784 (0.778, 0.789) | 0.967 (0.941, 0.993) | 0.007 (-0.024, 0.037) |
| | Stacked: 3-way poly | 8 | 0.784 (0.779, 0.789) | 1.004 (0.978, 1.030) | 0.009 (-0.021, 0.040) |
| | De novo | 31 | 0.803 (0.799, 0.808) | 0.952 (0.927, 0.976) | 0.016 (-0.015, 0.046) |
